# Actigraphy Informs Distinct Patient-Centered Outcomes in Pre-COPD

**DOI:** 10.1101/2021.05.01.21256454

**Authors:** Jianhong Chen, Lemlem Weldemichael, Brian Giang, Jeroen Geerts, Siyang Zeng, Wendy Czerina Ching, Melissa Nishihama, Warren M Gold, Mehrdad Arjomandi

**Author notes:** **Corresponding Author Information: Mehrdad Arjomandi, MD**, Division of Pulmonary and Critical Care Medicine, University of California San Francisco, San Francisco Veterans Affairs Medical Center, Bldg 203, Room 3A-128, Mailstop 111-D, 4150 Clement Street, San Francisco, CA 94121. These authors contributed equally to this work (Jianhong Chen, Lemlem Weldemichael). **Email Addresses:** JC.

## Abstract

**Background:** Actigraphy can clarify useful patient-centered outcomes for quantification of physical activity in the “real-world” setting.

**Methods:** To characterize the relationship of actigraphy outputs with “in-laboratory” measures of cardiopulmonary function and respiratory symptoms in pre-COPD, we obtained actigraphy data for 8 hours/day for 5 consecutive days a week before in-laboratory administration of respiratory questionnaires, PFT, and CPET to a subgroup of subjects participating in the larger study of the health effects of exposure to secondhand tobacco smoke who had air trapping but no spirometric obstruction (pre-COPD). Using machine learning approaches, we identified the most relevant actigraphy predictors and examined their associations with symptoms, lung function, and exercise outcomes.

**Results:** Sixty-one subjects (age=66±7years; BMI=24±3kg/m^2^; FEV_1_/FVC=0.75±0.05; FEV_1_=103±17%predicted) completed the nested study. In the hierarchical cluster analysis, the activity, distance, and energy domains of actigraphy, including moderate to vigorous physical activity, were closely correlated with each other, but were only loosely associated with spirometric and peak exercise measures of oxygen consumption, ventilation, oxygen-pulse, and anaerobic threshold (VO_2AT_), and were divergent from symptom measures. Conversely, the sedentary domain clustered with respiratory symptoms, air trapping, airflow indices, and ventilatory efficiency. In Regression modeling, sedentary domain was inversely associated with baseline lung volumes and tidal breathing at peak exercise, while the activity domains were associated with VO_2AT_. Respiratory symptoms and PFT data were not associated with actigraphy outcomes.

**Discussion:** Outpatient actigraphy can provide information for “real-world” patient-centered outcomes that are not captured by standardized respiratory questionnaires, lung function, or exercise testing. Actigraphy activity and sedentary domains inform of distinct outcomes.

**VISUAL ABSTRACT:** 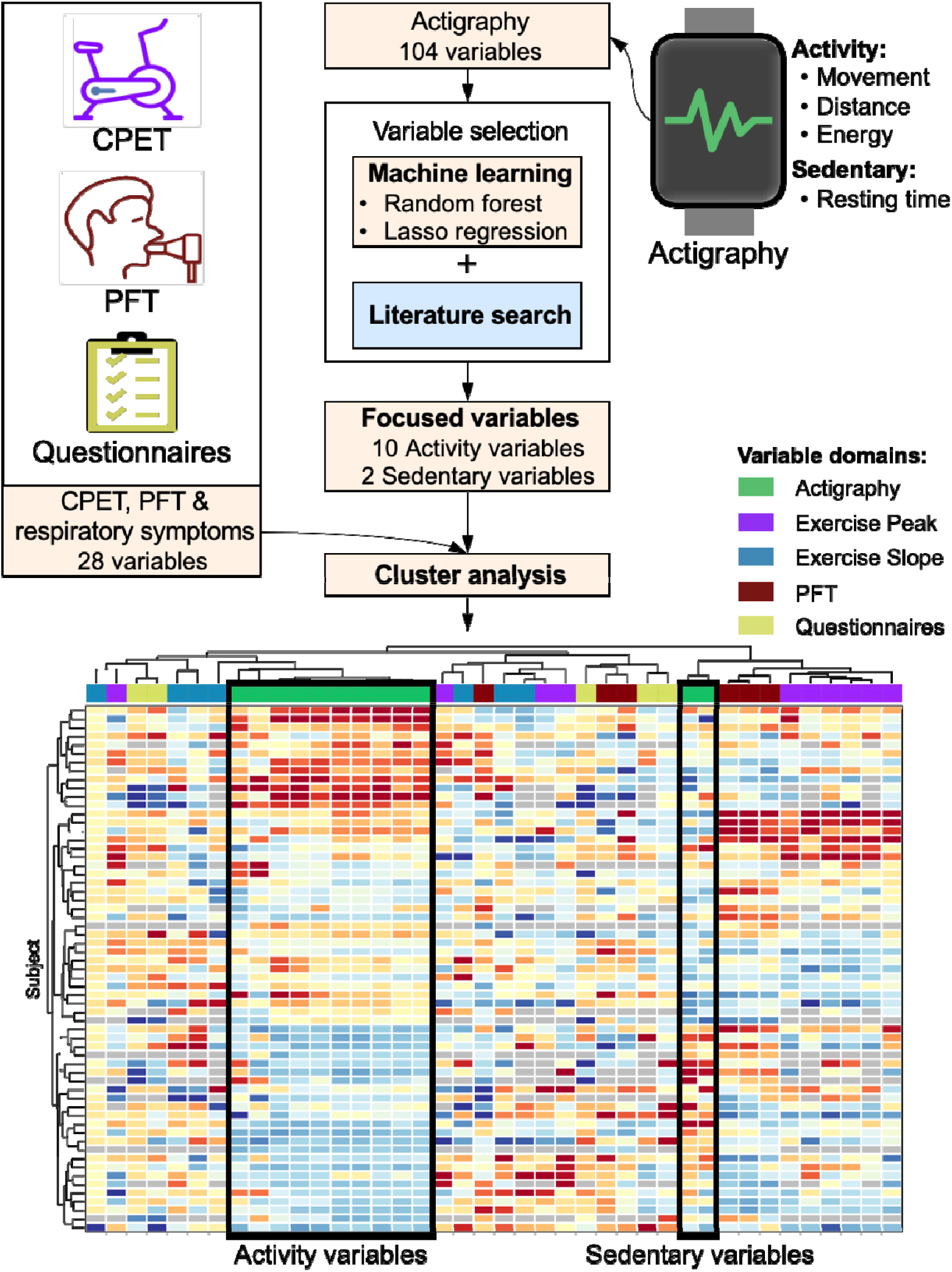

## INTRODUCTION

The ability to assess and quantify the physical activity levels of patients can offer valuable insights into their general health and day-to-day functioning that may not be captured by traditional objective measurements of disease status, such as interview- or questionnaire-based evaluations or physiological or laboratory testing.^1-3^ Wearable activity monitors that measure “real-world” physical activity levels may thus serve as objective and reliable tools to assess health status and disease activity.^4-7^ The current-generation wearable activity monitors, including pedometers and accelerometers, are transforming the field of biomedical research by their capacity to approximate free-living conditions and measure real-world physical activity in a continuous and longitudinal, yet objective manner.^8^ However, the correlation between physical activity measurements obtained by actigraphy and the physical activity outcomes measured by physiologic and questionnaire-based tools has not been clearly identified.

In this study, we aimed to understand the relationship between physical activity measures obtained from a wearable activity monitor and those obtained from standardized respiratory questionnaires and “in-laboratory” lung function and exercise testing. We hypothesized that “real-world” actigraphy provides distinct outcomes that are not completely captured by standardized symptom questionnaires or traditional “in-laboratory” functional assessments. To assess this hypothesis, we examined the association of outpatient actigraphy measures with patient-reported respiratory physical activity and symptoms as well as lung function and exercise test results in a never-smoker cohort subgroup of a study on the health effects of exposure to secondhand tobacco smoke (SHS); the participants were at risk for COPD due to their prolonged occupational exposure to SHS but showed preserved spirometry.

## METHODS

### Study overview

This observational study was nested in a larger study examining the cardiopulmonary health effects of SHS exposure in a cohort of nonsmoking individuals with a range of occupational SHS exposure, as previously described.^9^ Briefly, between July 2007 and March 2020, we recruited US airline flight crewmembers with a history of occupational exposure to SHS, along with nonsmoker controls without such occupational exposure, who were participating in a larger study of the cardiopulmonary health effects of prolonged exposure to SHS (ClinicalTrials.gov Identifier: NCT02797275). The participants underwent respiratory symptom questionnaire assessments, full pulmonary function testing (PFT), and maximum-effort cardiopulmonary exercise testing (CPET). For the actigraphy nested study, beginning February 2014, participants were asked to wear an activity monitor eight hours a day for five consecutive days during the week before they came in for in-laboratory evaluation and maintain a daily diary. The actigraphy data were then obtained and analyzed along with the respiratory questionnaire, PFT, and CPET data to examine its association with reported physical activity, symptoms, and in-laboratory measures of physical activity. The University of California San Francisco (UCSF) Institutional Review Board (IRB) and the San Francisco VA Medical Center Committee on Research and Development approved the study protocols. Full details of the methods are available in the **Supplemental Appendix**.

### Study Population

US airline crewmembers, including flight attendants and pilots, were eligible to participate if they had worked for ≥5 years in an aircraft. A reference group of “sea-level” participants who lived in San Francisco Bay area and had never been employed as flight crewmembers was also recruited. Participants were eligible if they were nonsmokers defined by never-smoking or, in ever smokers, no smoking for ≥20 years and a cumulative smoking history of <20 pack-years. Eligible participants were excluded if they had a known history of cardiopulmonary disease or recreational drug use, including marijuana consumption. All participants enrolled in the larger cohort were invited to participate in this nested study. For the nested actigraphy study, recruitment began in February 5, 2014 and continued through March 17, 2020.

### Physical Activity Monitoring using Actigraphy

Physical activity was monitored using a triaxial accelerometer-based activity monitor (ActiGraph GT3X; Actigraph Corporations, Pensacola, FL). Technical details of the device can be found in the **Supplemental Appendix**. The ActiGraph monitor was initialized to continuously collect data over a period of 5 days. It was then mailed to participants along with a daily diary to keep a log of the time the monitor was worn and the activities the participants performed during that time. All participants received the ActiGraph monitors at least 7 days prior to the in-laboratory visit, during which respiratory symptom questionnaire, PFT, and CPET data were collected. Participants were instructed to wear the ActiGraph monitor on the waist upon awakening using a wrist band provided and to keep it on continuously for at least 8 hours for 5 consecutive days beginning the start of the first day of their work week. The 5-day monitoring period was chosen to allow for adequate weekday data collection^10^ while avoiding recording of non-routine rest or activity periods that typically occur during weekends. All participants were carefully instructed on correctly positioning the device.

### Actigraphy Data Description and Processing

Actigraphy data were processed using the ActiLife software program (Version 6.11.9; ActiGraph LLC) and saved in raw format as GT3X files. The ActiLife software generates a total of 52 variables in the distance, time (activity and sedentary), and energy domains. The list of variables and their definitions are shown in **Table S1**. The actigraphy data were matched against the diary to ascertain appropriate usage of the monitor, and the data were considered to be acceptable if the participants wore the monitor for a minimum of 3 days and for more than 5 hours (300 min) per day. The data was summarized into 5-day average values (weekly “mean” values) and the highest values of “maximum” values (maximum per epoch) across all 5 days (weekly “highest” values). The total amount of the time that the monitor was worn was included in the regression models as *total time worn*.

### Actigraphy Data Variable Selection

Actigraphy generates a large number of variables, some of which are highly correlated. Given the low number of participants in our study, we decided to reduce the number of actigraphy variables to increase the robustness of the analysis. To achieve this, we pursued two approaches of variable selection: (1) machine learning approach, and (2) literature-guided approach (**Figure 1**).

**Figure 1.**
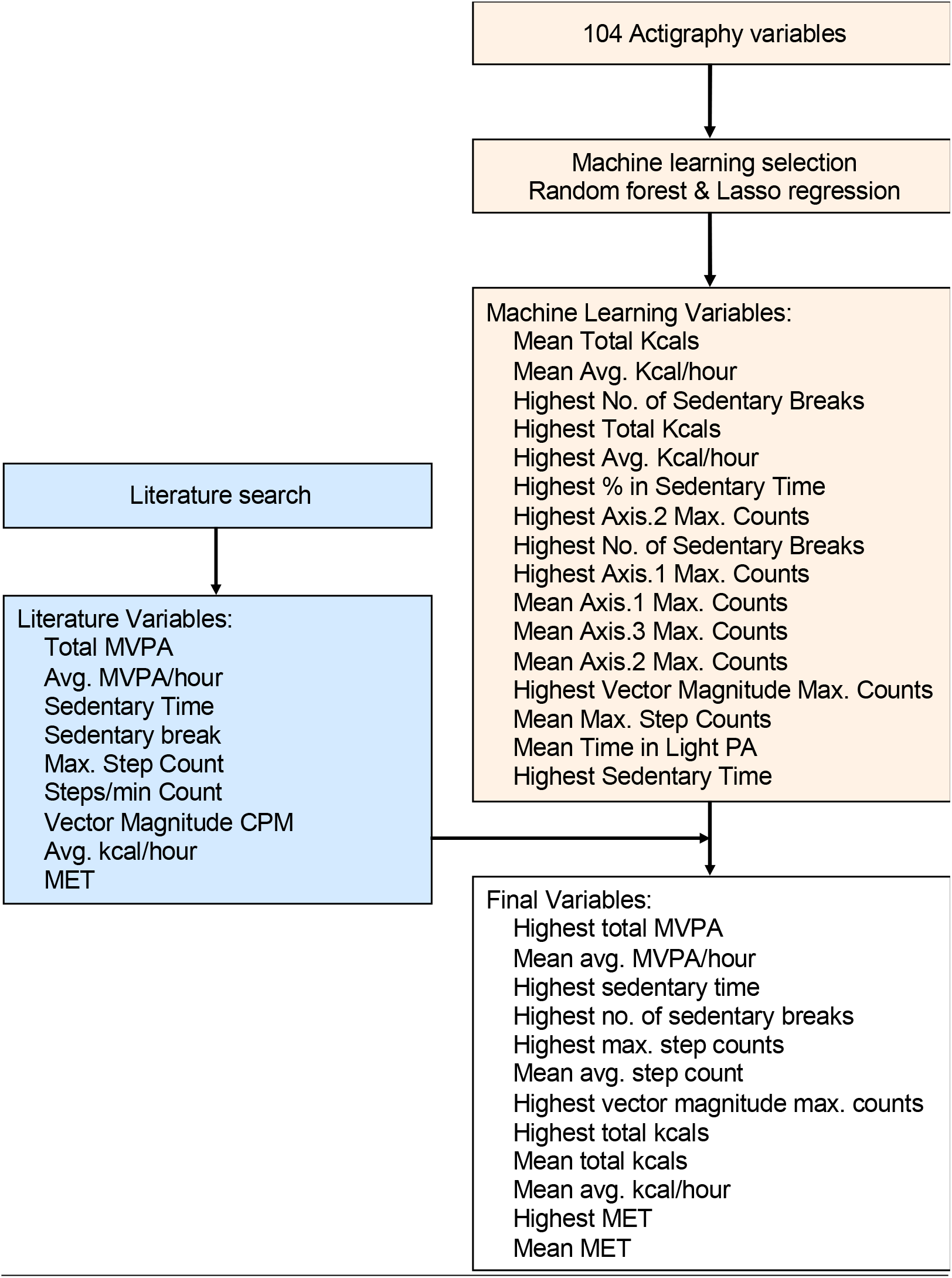
Actigraphy variable selection. A schematic to show the process of variable selection by combining machine learning and literature guided approaches. Machine learning approach was done by implementing random forest and lasso regression. The top ranked 10 variables from each method were done used against a list of variables extracted from the available literature, and a final “focused” set of variables representing all actigraphy domains were selected. “Highest” and “Mean” for actigraphy variables represent the highest and mean of 5-day activity monitoring measurements.

In the machine learning approach, we built two models, random forest and lasso regression. Using random forest modeling, we generated a list of top-ranked actigraphy variables that were predictive of various questionnaire, PFT, and CPET outcomes by ranking variables based on minimizing prediction error at the splitting of the decision tree nodes. For lasso regression, we used a similar strategy to generate a list of important variables by utilizing the l_1_- norm penalized terms to force unimportant variables to become zero. To rank the variables in lasso regression, we then compared the magnitude of variable coefficients in various models. Finally, we summarized the actigraphy variables by including the top 10 variables from each model.

In the literature-guided approach, we reviewed the available literature on actigraphy and selected 9 variables to represent distance, energy, and the activity and sedentary time domains as described below (**Table S2**). The final set of variables was selected based on a combination of the machine learning and literature-guided approaches to provide meaningful variables of highest predictive value for our proposed analysis.

### Pulmonary Function and Cardiopulmonary Exercise Testing

Details of our PFT and CPET procedures are presented in the **Supplemental Appendix** and have been previously described.^9,11,12^

### Respiratory Questionnaires

Patient-reported respiratory symptoms, physical activity, and quality of life assessments were conducted using the COPD Assessment Test (CAT),^13^ modified Medical Research Council (mMRC) Dyspnea Scale,^14^ the Short Form 12-Item Health Survey (SF-12),^15^ International Physical Activity Questionnaire (IPAQ),^16^ and Airway Questionnaire 20 (AQ20).^17^

### Data Analysis

Distributions of patients’ actigraphy, respiratory symptom, lung function, and exercise data were visualized and inspected (**Figure S1**). Because most variables were not normally distributed, Spearman’s rank correlation was used throughout the analysis. We then examined the association of actigraphy distance, energy, and activity and sedentary time domains with respiratory questionnaire, PFT, and CPET outputs after adjustment for age, sex, height, weight, and time worn using hierarchical clustering with the Spearman correlation coefficient as the distance metric.

Even after selection of a focused set of actigraphy variables, the initial exploratory analyses showed two potential statistical challenges, namely, high dimensionality and high collinearity, for the application of these variables to ordinary linear regression modeling. To address these challenges, we employed partial principal component regression, which combined principal component analysis with adjustment for covariates to reduce the number of dimensions and transformed the original variables into orthogonal principal component (PC) axes. Next, we performed ordinary linear regression using the top six PC axes as the new predictive actigraphy variables within the model. We later computed the Pearson correlation coefficients between the original actigraphy variables versus the transformed PC axes to help interpret the representation of each PC axis. We used P-values of 0.05 as the cutoff for statistical significance and considered Spearman and Pearson correlation coefficients ±0.5 as indicating strong correlation (either positive or negative).

## RESULTS

### Participant Characteristics

Overall, 64 volunteers participated in the nested study and wore the actigraphy monitor during the week before participating in the in-laboratory assessments with respiratory questionnaires, PFT, and CPET. Three of the 64 subjects wore the actigraphy monitor for <3 consecutive days or <300 consecutive min/day and thus were excluded from the analysis. The characteristics of the 61 participants included in the analysis are shown in **Table 1**. The participants were aged 65.7±10.7 years, predominantly female (57 [93%]), and never-smokers. All participants had a history of occupational exposure to SHS through their airline employment as flight crew for a median (interquartile range, [IQR]) period of 15.6 (6.0-25.0) years. All participants showed preserved spirometry, with forced expiratory volume in 1 s (FEV_1_) and FEV_1_/forced vital capacity (FVC) percent predicted values of 103±17 and 98±6, respectively. All of them showed mild air trapping, as defined by residual volume (RV)/total lung capacity (TLC) > 0.35 (0.38±0.06), and an overall reduced average diffusing capacity adjusted for hemoglobin of 67%±28% predicted. The percent predicted values for the measures of VO_2_, V_E_, and oxygen-pulse at the peak of exercise (VO_2Peak_, V_EPeak_, and O_2_-PulsePeak) were 103.8±18.1, 61.1±14.3, and 109.4±20.7, respectively, and VO_2_ at anaerobic threshold (VO_2AT_) was 63.8%±16.7% of VO_2Peak_. **Table 1** also shows the selected actigraphy data in the distance, energy, and activity and sedentary time domains. Overall, the participants wore the actigraphy monitor for a median (IQR) period of 5 (4-5) days and 38.9 (33.0-39.9) h (2,334 [1,980-2,396] min).

**Table 1.**
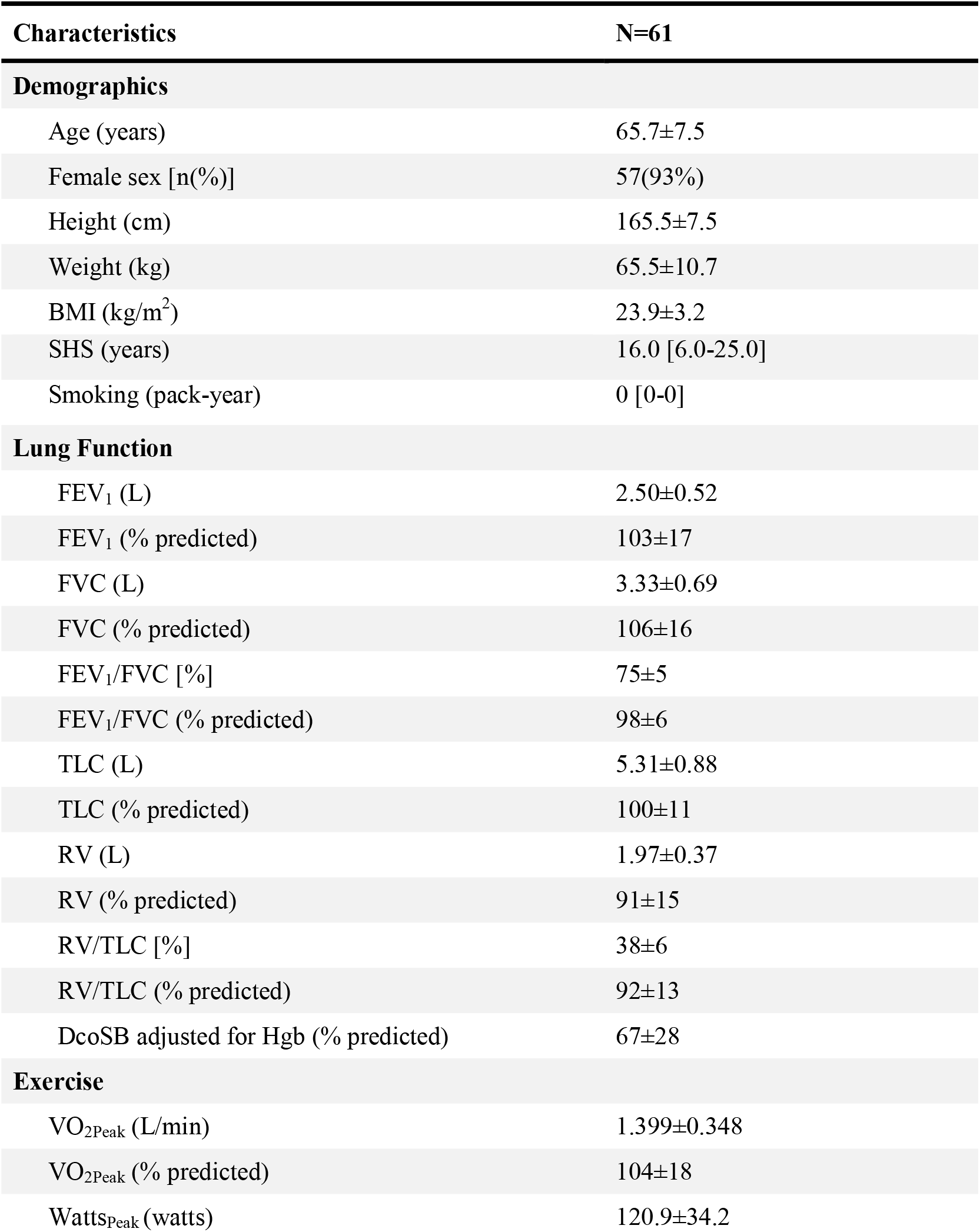
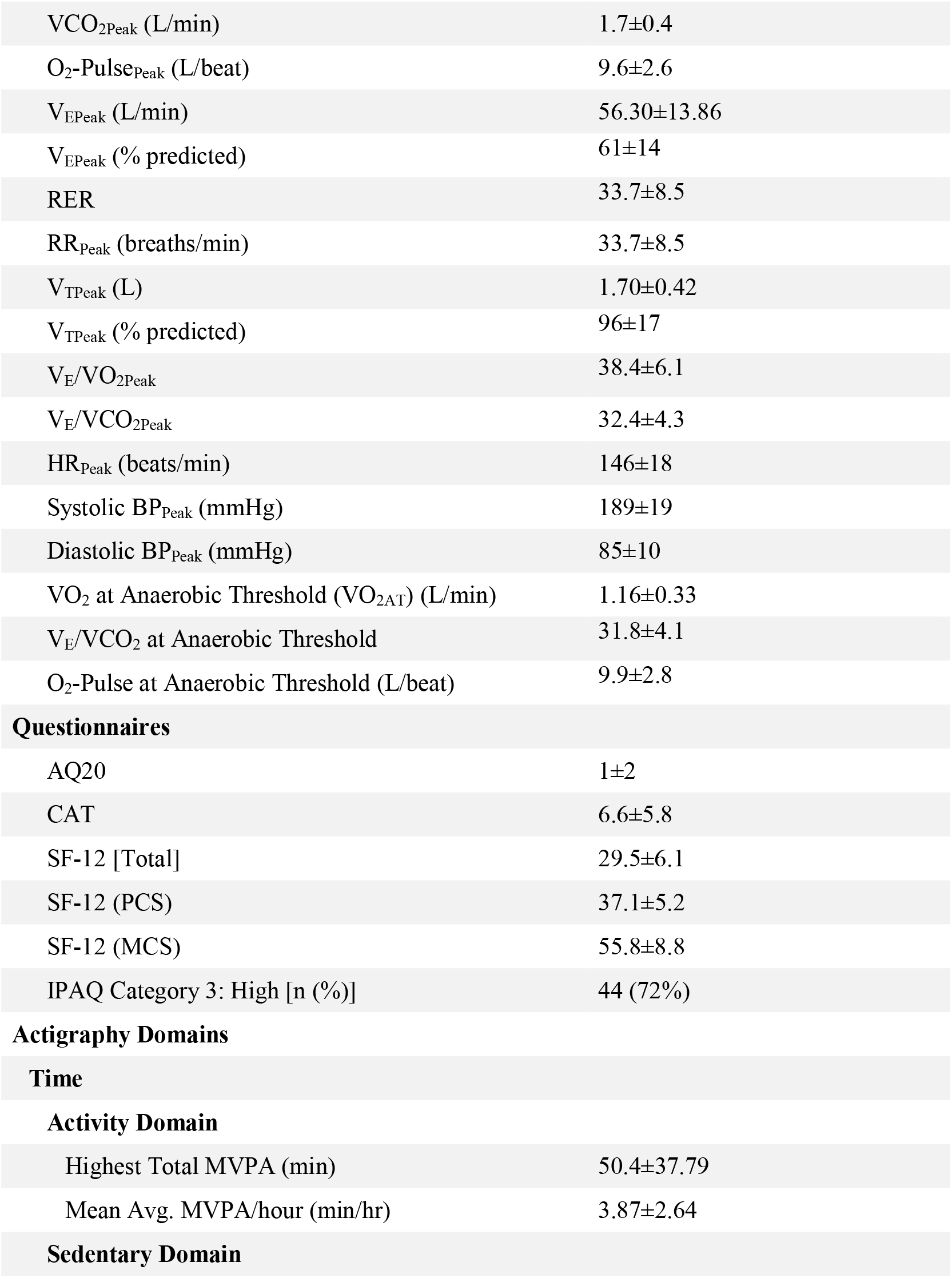
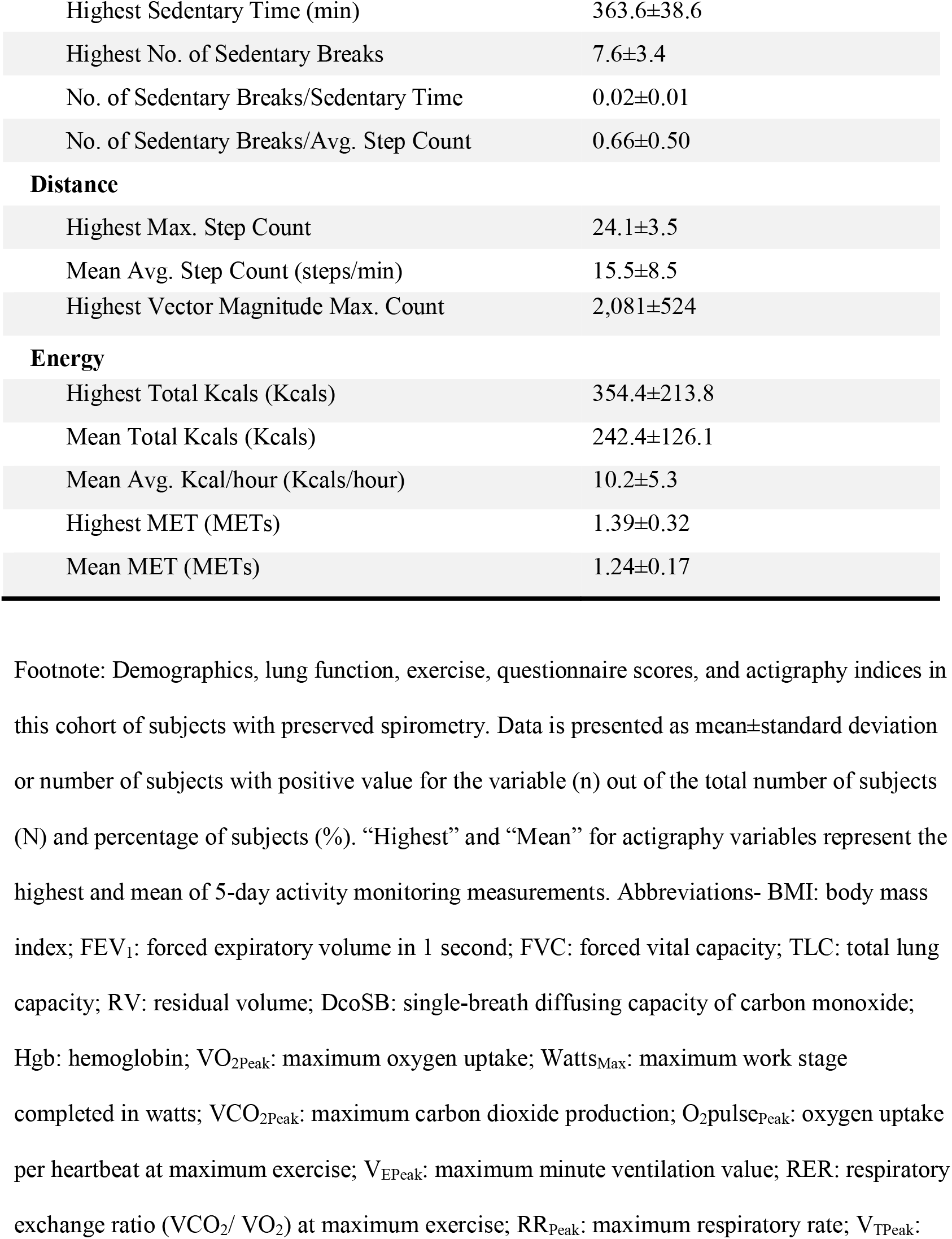
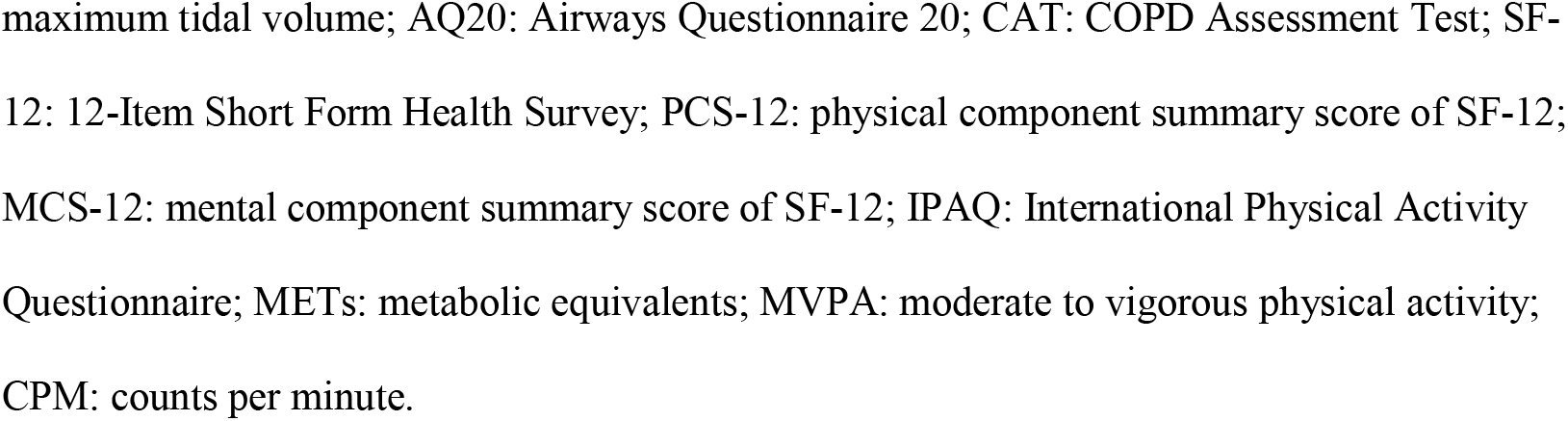
Subject characteristics.

### Combined Approach for Selection of Appropriate Actigraphy Variables

In the machine learning approach, random forest and lasso regression analyses were performed to identify the most relevant actigraphy variables that are predictive of the respiratory questionnaire, PFT, and CPET outcomes. The top-ranked variables for each machine learning approach, which are shown in **Figure S2 A and B**, were a mix of actigraphy domains, including the sedentary time domain. The ten top ranked variables from each approach were then matched against a list of most highly cited actigraphy variables in the literature, and a final set of actigraphy variables, which included variables from all domains, were then generated for further analysis (**Figure 1**). The final set of variables obtained by this combined approach is shown in **Table 2**.

**Table 2.**
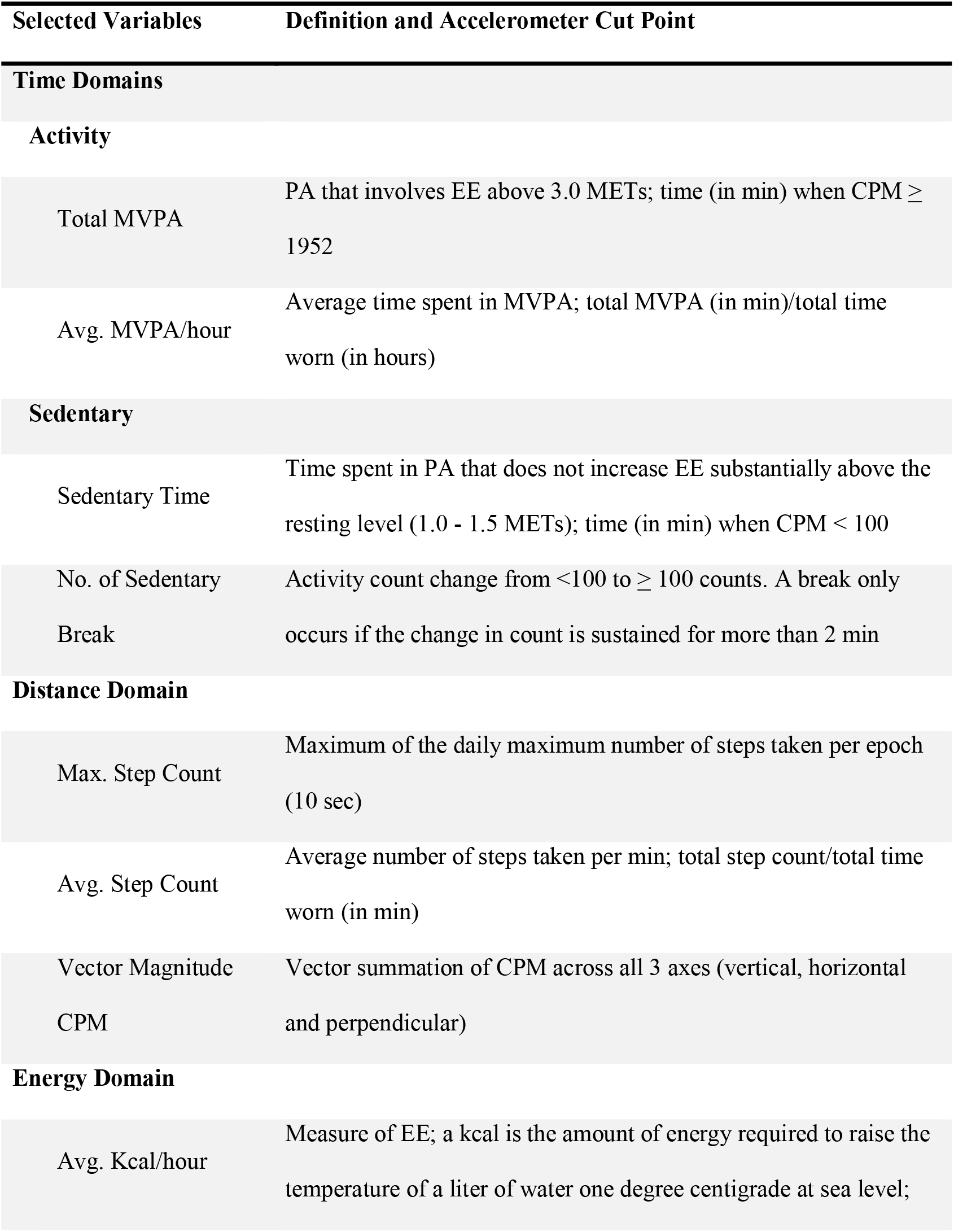
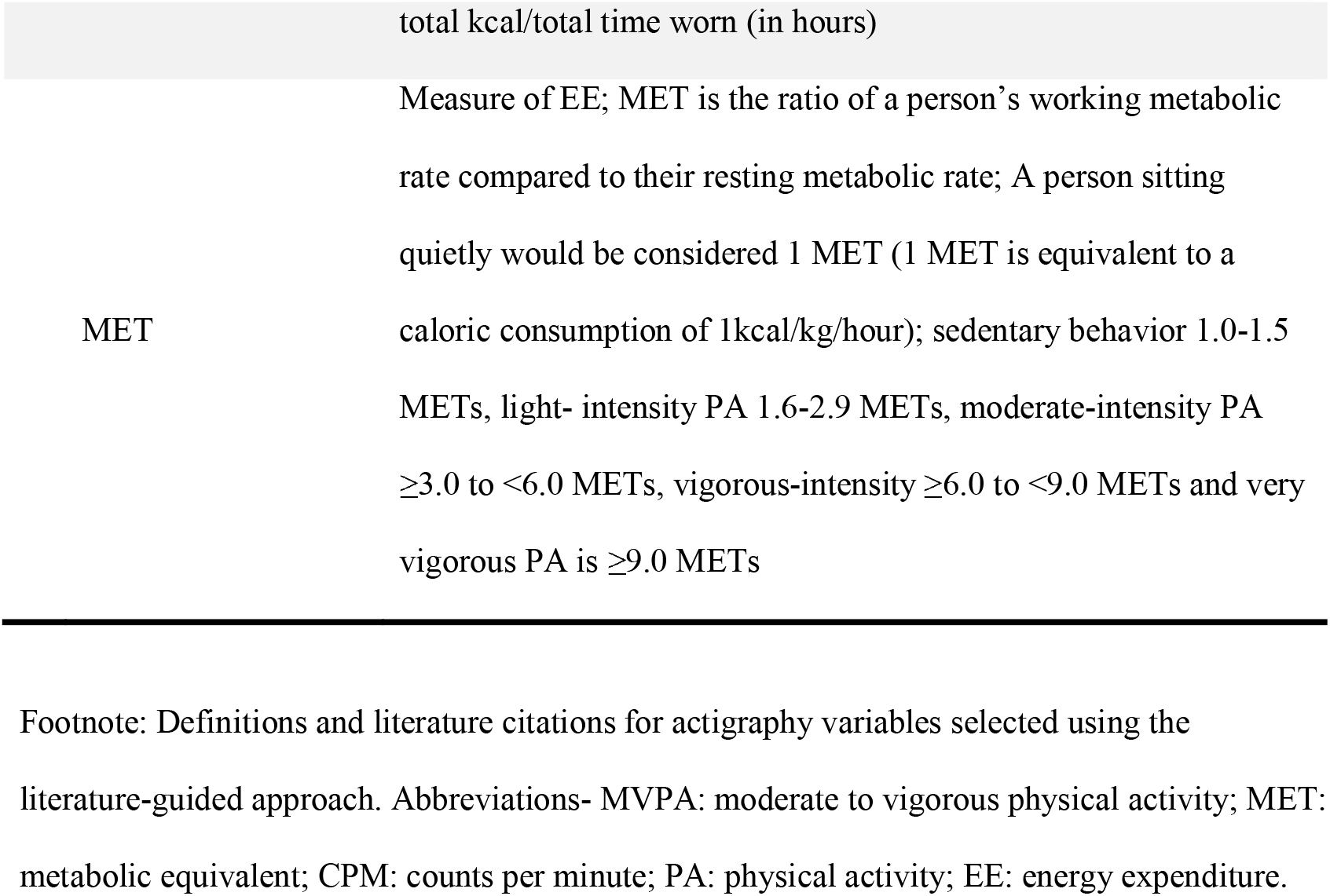
Definitions of final selection of actigraphy variables.

The correlations among all and the final selection of actigraphy variables are shown in **Figure S1** and **Figure 2**, respectively. Variables from the distance, energy, and activity time domains were highly and directly correlated with each other, with the strongest correlation observed between the energy and activity time domains and the weakest between these domains and the distance domain. However, sedentary time domain variables were poorly and inversely correlated with the variables from the distance, energy, and activity time domains (**Figure 2**). Within the sedentary time domain, the total amount of time spent in the sedentary condition (Sedentary Time) was closely and directly associated with the number of times participants broke their sedentary condition (No. of Sedentary Breaks; r=0.65; P<0.001).

**Figure 2.**
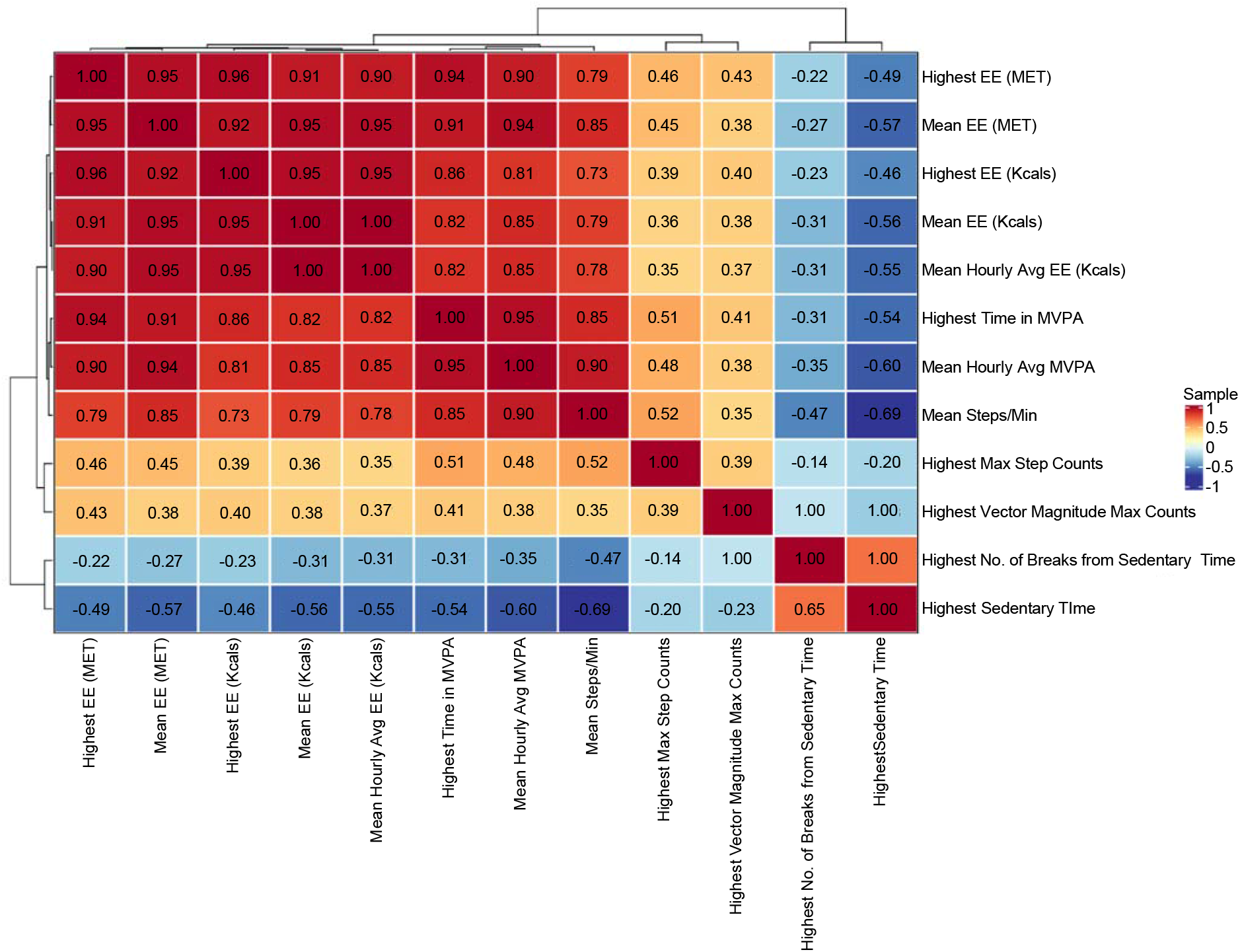
Correlation among final set of actigraphy variables. Spearman correlation coefficient in clustering heat map of the final selection of actigraphy variables. Visualization shows that almost all of actigraphy variables are highly correlated.

### Hierarchical Clustering Identified Two Clusters of Actigraphy Domains That Distinctly Group with Respiratory Questionnaire, PFT, and CPET Outcomes

Hierarchical clustering analysis categorized actigraphy domains into two distinct clusters (**Figure S3** and **Figure 3**). Measures of the distance, energy, and activity time domains, including moderate-to-vigorous physical activity (MVPA), maximum number of steps taken per epoch (Max. Step Count), and average hourly energy expenditure (Avg. Kcal/hour), clustered together and were best correlated with the rate of increase in O_2_-Pulse, tidal breathing, systolic blood pressure over workload (O_2_-Pulse_Slope_, SBP_Slope_, and V_TSlope_,), and SF-12 and its “physical” component score, and to a lesser extent with air trapping (RV and RV/TLC), airflow obstruction (FEV_1_/FVC), ventilatory efficiency at peak exercise (V_E_/VCO_2Peak_), and other respiratory questionnaires (AQ-20 and CAT) scores. Conversely, measures of the sedentary domain, including Sedentary Time and No. of Sedentary Breaks, clustered together with spirometric indices (FEV_1_, FVC), TLC, and peak exercise measures of oxygen consumption (VO_2Peak_), ventilation (V_EPeak_), tidal volume (V_TPeak_), and oxygen-pulse (O_2_-Pulse_Peak_).

**Figure 3.**
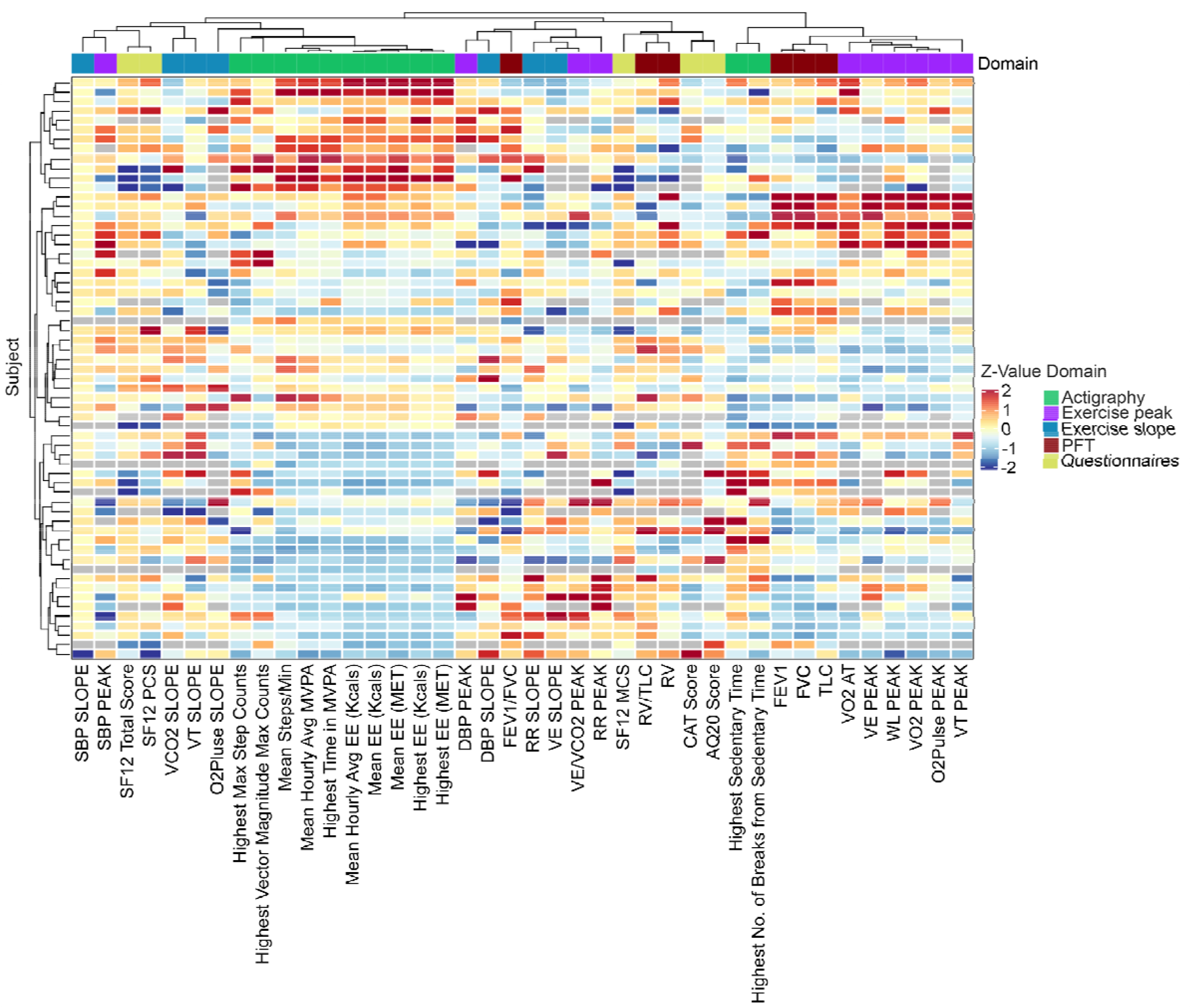
Hierarchical clustering analysis and heatmap representation of final selection of actigraphy variables with questionnaire, PFT, and CPET measures. We performed cluster analysis using Spearman correlation coefficients as the distance metric. The clustered relationships were present as the dendrograms on the top and left size of the heat map. Two distinct clusters of actigraphy variables were grouped together from the three domains (time, distance, energy). The two clusters were based on activity related (such as total energy spent in Kcals, number of steps walked, etc.) and sedentary related (such as sedentary time, break, etc.) respectively. Each row represents one subject. Grey box are missing values.

### Principal Component Analysis of a Focused Dataset of Actigraphy Variables Reproduced Contrasting Actigraphy Domains Represented by Two Principal Components

To address the high dimensionality and collinearity of actigraphy variables, we performed principal component analysis (**Table 3**) and regression of the final set of variables (**Table 4**). The first six principal components (PC) explained 87.8% of the variation in the dataset and were chosen for further analysis. The most contributing PC (PC1) showed high positive values for the following parameters: measures of energy expenditure (Avg. Kcal/hour=0.92, Highest MET=0.94, and Mean MET=0.96), maximum number of steps taken per epoch (Max. Step Count=0.56), average number of steps taken per minute (Avg. Steps/min=0.91), maximum number of steps counts on actigraphy axes 1 and 3, and as a vector magnitude across all 3 axes (Vector Magnitude Max. Count=0.4), time spent in MVPA (Highest Total MVPA=0.92; Avg. MVPA/hour=0.94). Therefore, PC1 was determined to be a reflection of the measures of activity time, distance, and energy domains of actigraphy. The second PC (PC2) was highly correlated with participant sex (0.77), height (0.84), and weight (0.84). The third PC (PC3) was highly correlated with the total time spent in the sedentary condition (Highest Sedentary Time=0.66), and thus was determined to be representative of the sedentary time domain of actigraphy. PC4 was only highly correlated with subject’s age (0.77).

**Table 3.**
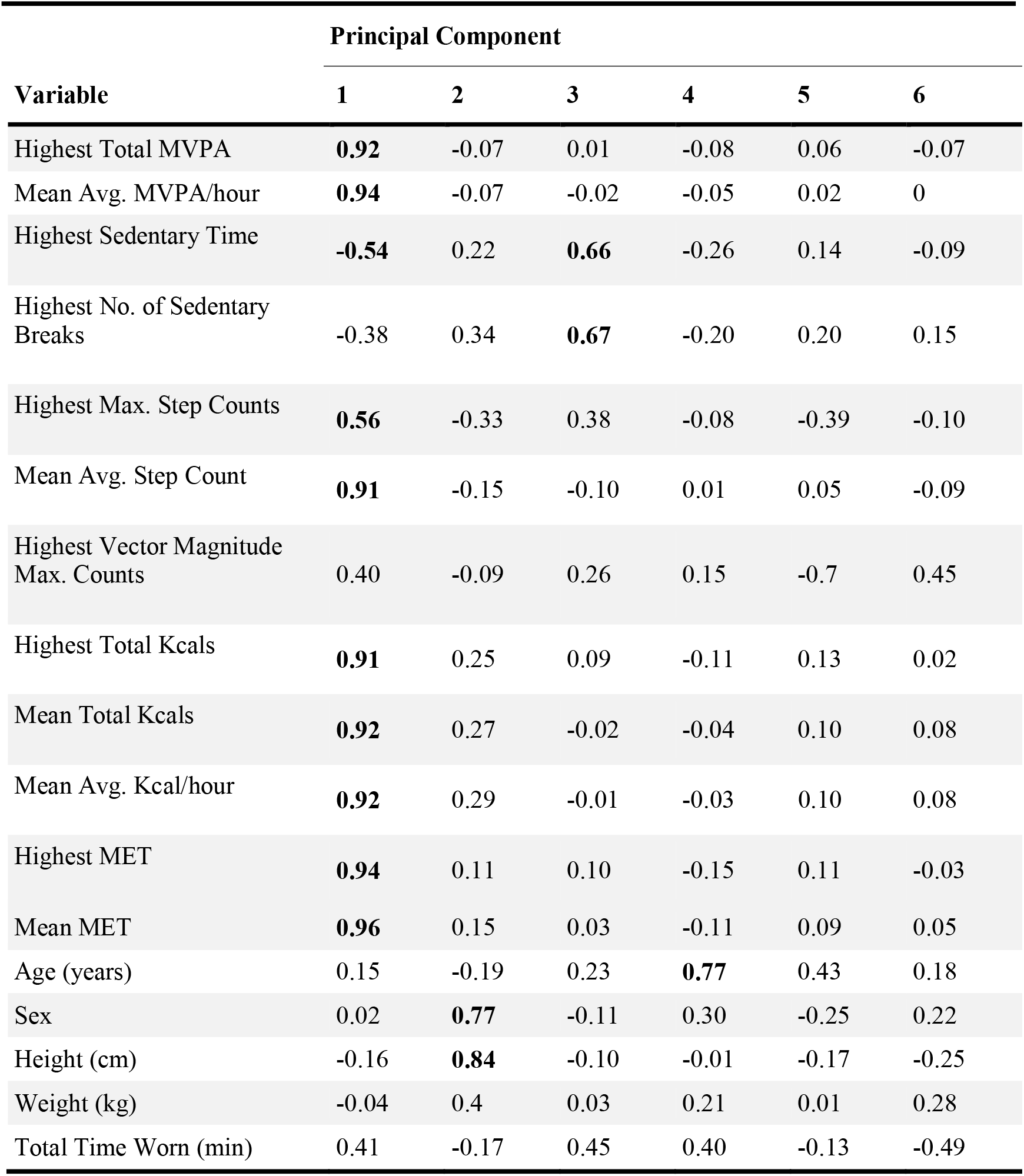
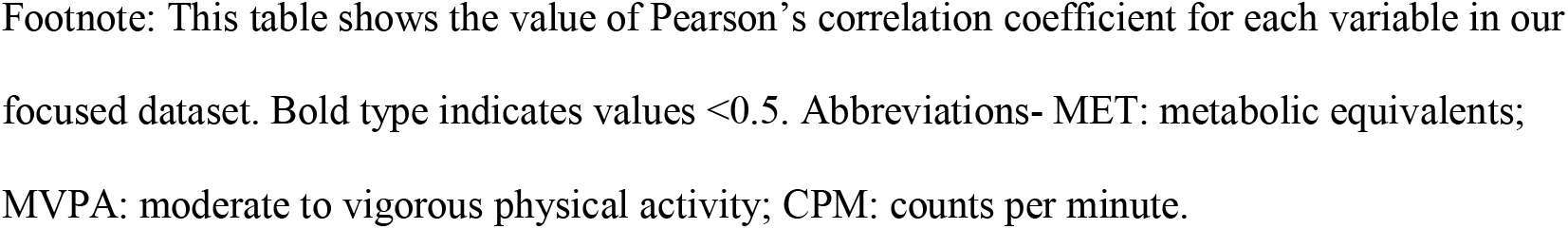
Results of the principal component analysis.

**Table 4.** Results of principal component regression analysis.

| Outcomes | PC1 (activity component) |  |  |  | PC3 (sedentary component) |  |  |  | Model Error |
| --- | --- | --- | --- | --- | --- | --- | --- | --- | --- |
|  | Estimate | Lower CI | Upper CI | P-value | Estimate | Lower CI | Upper CI | P-value | RMSE |
| **Lung function** |  |  |  |  |  |  |  |  |  |
| FEV<sub>1</sub> (L) | 0.005 | -0.032 | 0.042 | 0.808 | **-0.094** | **-0.183** | **-0.006** | **0.042** | 0.39 |
| FVC (L) | -0.002 | -0.050 | 0.046 | 0.924 | -0.107 | -0.222 | 0.009 | 0.075 | 0.51 |
| FEV<sub>1</sub>/FVC [%] | 0.002 | -0.002 | 0.007 | 0.284 | -0.004 | -0.014 | 0.006 | 0.458 | 0.05 |
| RV (L) | -0.005 | -0.037 | 0.028 | 0.781 | -0.045 | -0.131 | 0.04 | 0.305 | 0.32 |
| TLC (L) | -0.016 | -0.077 | 0.044 | 0.597 | -0.143 | -0.288 | 0.001 | 0.057 | 0.32 |
| RV/TLC [%] | -0.001 | -0.006 | 0.004 | 0.796 | 0.003 | -0.010 | 0.017 | 0.617 | 0.05 |
| **Exercise** |  |  |  |  |  |  |  |  |  |
| VO<sub>2Peak</sub> (L/min) | 0.002 | -0.020 | 0.025 | 0.838 | 0.016 | -0.039 | 0.070 | 0.575 | 0.23 |
| O<sub>2</sub>-Pulse<sub>Peak</sub> (L/beat) | 0.179 | -0.021 | 0.379 | 0.088 | 0.131 | -0.327 | 0.590 | 0.578 | 1.52 |
| V<sub>EPeak</sub> (L/min) | 0.133 | -1.217 | 1.482 | 0.848 | 0.231 | -3.144 | 3.607 | 0.894 | 11.25 |
| RR<sub>Peak</sub> (breaths/min) | -0.611 | -1.390 | 0.167 | 0.130 | 1.741 | -0.171 | 3.654 | 0.081 | 7.92 |
| V<sub>TPeak</sub> (L) | 0.006 | -0.022 | 0.035 | 0.660 | -0.062 | -0.131 | 0.008 | 0.087 | 0.29 |
| V<sub>E</sub>/VCO<sub>2Peak</sub> | -0.147 | -0.625 | 0.331 | 0.551 | 0.314 | -0.882 | 1.509 | 0.610 | 3.99 |
| Watts<sub>Peak</sub> (watts) | 1.067 | -0.939 | 3.072 | 0.302 | -0.104 | -5.029 | 4.821 | 0.967 | 20.40 |
| Systolic BP<sub>Peak</sub> (mmHg) | 0.112 | -1.639 | 1.863 | 0.901 | -0.376 | -4.696 | 3.945 | 0.865 | 17.51 |
| Diastolic BP<sub>Peak</sub> (mmHg) | 0.515 | -0.488 | 1.518 | 0.320 | 0.042 | -2.434 | 2.518 | 0.974 | 10.04 |
| VO<sub>2</sub> at Anaerobic Threshold (L/min) | **0.032** | **0.005** | **0.059** | **0.026** | 0.005 | -0.065 | 0.076 | 0.885 | 0.25 |
| VCO<sub>2Slope</sub> (L/min) | -0.005 | -0.017 | 0.007 | 0.405 | -0.002 | -0.031 | 0.028 | 0.917 | 0.12 |
| O<sub>2</sub>-Pulse<sub>Slope</sub> (L/beat) | 0.071 | -0.016 | 0.158 | 0.118 | 0.127 | -0.090 | 0.344 | 0.259 | 0.73 |
| V<sub>ESlope</sub> (L/min) | -0.516 | -1.163 | 0.132 | 0.127 | 0.185 | -1.435 | 1.805 | 0.824 | 5.40 |
| RR<sub>Slope</sub> (breaths/min) | -0.183 | -0.714 | 0.349 | 0.503 | 0.128 | -1.177 | 1.1434 | 0.848 | 5.41 |
| V<sub>TSlope</sub> (L) | -0.016 | -0.046 | 0.015 | 0.315 | -0.025 | -0.099 | 0.049 | 0.507 | 0.31 |
| Systolic BP<sub>Slope</sub> (mmHg) | 8.095 | -9.451 | 25.641 | 0.370 | 18.368 | -24.726 | 61.462 | 0.408 | 178.45 |
| Diastolic BP<sub>Slope</sub> (mmHg) | 0.205 | -0.768 | 1.177 | 0.682 | -0.936 | -3.325 | 1.453 | 0.446 | 9.89 |
| **Questionnaires** |  |  |  |  |  |  |  |  |  |
| AQ20 | -0.150 | -0.336 | 0.036 | 0.120 | 0.261 | -0.193 | 0.715 | 0.266 | 1.86 |
| CAT | -0.332 | -0.980 | 0.316 | 0.322 | -0.101 | -1.697 | 1.496 | 0.902 | 5.31 |
| SF-12 [Total] | -0.319 | -0.880 | 0.241 | 0.270 | **-1.471** | **-2.840** | **-0.103** | **0.041** | 5.60 |
| SF-12 (PCS) | -0.026 | -0.513 | 0.462 | 0.918 | -0.451 | -1.641 | 0.738 | 0.461 | 4.87 |
| SF-12 (MCS) | -0.791 | -1.568 | -0.014 | 0.052 | **-2.102** | **-3.999** | **-0.204** | **0.035** | 7.76 |

Results of the principal component regressions of the cardiopulmonary function and questionnaires outcomes over PC1 (“activity component) and PC3 (“sedentary component”) are presented in **Table 4**. In the activity domains represented by PC1, a significant positive association was only observed with the volume of oxygen consumption at anaerobic threshold (VO_2AT_) (P=0.026). In the actigraphy sedentary domain represented by PC3, significant negative associations were observed with FEV_1_ (P=0.042) (marginally non-significance with FVC [P=0.075] and TLC [P=0.057]) and the total and “mental” component of SF-12 score (P=0.042 and P=0.035, respectively). No other significant associations were present.

## DISCUSSION

In this observational study of a population at risk for COPD due to prolonged occupational exposure to SHS but with no airflow obstruction (“pre-COPD”), we employed a combined machine learning and literature-based approach to tackle the high dimensionality of the actigraphy output variables and identified a set of “focused” variables that were most relevant to and best represented the daily physical activity pattern of subjects. We then pursued two different analytical strategies. In the first strategy, we used unsupervised cluster analysis to evaluate the general relationship among the in-laboratory physiological measures and questionnaire-based respiratory symptom scores with the focused set of actigraphy variables using an unadjusted approach and without any assumption of linearity to show how variables from different modalities may correlate with each other. Using this strategy, we found sedentary domain of actigraphy to be more closely grouped together with exercise variables representing lower peak exercise capacity and lack of anaerobic fitness, while distance, activity, and energy domains of actigraphy were more closely grouped together with variables representing presence of endurance and aerobic fitness (slope of variables over workload), better scores on respiratory questionnaires, and lung function indices of air trapping. In the second strategy, we used principal component regression to more exactly examine the associations between specific in-laboratory measures or specific respiratory symptom scores and focused actigraphy variables with adjustment for covariates in the models that assumed linear relationships. Using this strategy, we found only few significant associations. Although this overall lack of statistically significant associations could be due to the sample size examined, the general trends were consistent with the unsupervised cluster analysis strategy, and suggested that, at least in this population with pre-COPD, actigraphy and in-laboratory physiological assessment and respiratory questionnaires are at best poorly correlated.

Overall, in this population at risk for COPD due to prolonged occupational exposure to SHS but with no airflow obstruction, actigraphy seems to provide distinct information about daily physical activity and functionality that are not captured by in-laboratory lung function, exercise testing, or standardized respiratory questionnaires. A possible explanation for this observation is that in-laboratory physical activity assessments may provide an estimate of the maximum functional *capacity*, while actigraphy may be more informative of the real-world level of physical activity that represents the functional *status*. Although functional capacity represents an individual’s maximum capacity to perform physical activity, functional status may best represent the activities people actually do during the course of their daily lives,^18,19^ and in this population with pre-COPD, while the functional capacity may be preserved, the functional status is reduced, resulting in poor correlation. In past studies of this cohort, we have demonstrated the cohort to have abnormal diffusing capacity that was associated with lower exercise tolerance despite having preserved spirometry.^11,12^ In the same cohort and also another cohort of direct smokers with preserved spirometry (those with Global Initiative for Obstructive Lung Disease [GOLD] 0 category), we have shown that air trapping, as manifested by abnormal lung volumes (RV/TLC), could be present and is predictive of lower exercise tolerance.^9,20^ On the other hand, patients with overt COPD have airflow obstruction in addition to any potential abnormal diffusing capacity or lung volumes (air trapping) that they may experience. This airflow obstruction may reduce exercise capacity more remarkably and affect functional status, causing those with overt COPD to have a physical activity status that matches their maximum capacity. Corroborating this hypothesis are the reports that in overt COPD, activity monitor outputs have predictive value in the assessment of disease severity, exercise capacity, and healthcare utilization patients.^21-23^ Thus, it is possible that the association of actigraphy with in-laboratory physiological assessment and respiratory symptoms is varied across COPD disease spectrum and improves as disease severity worsens. These findings also implicate measurement of physical activity status by actigraphy as a useful tool for detection of early disease in COPD when the maximum functional capacity, measured by in-laboratory testing, may be less affected. Further research involving larger cohorts and across the entire COPD disease spectrum would be needed to better understand potential diagnostic application of actigraphy for assessment of patients’ real-world physical status.

Another intriguing finding in our study was that both unsupervised cluster analysis and principal component regression approaches showed that actigraphy variables to group into two distinct clusters of (1) distance, energy, and activity time domains and (2) sedentary time domain. Remarkably, these clusters did not show a simple inverse relationship, were rather poorly correlated, and grouped together differently with various measures of in-laboratory cardiopulmonary functional assessment and self-reported symptomatology, indicating that they are influenced by distinct physiological processes. These features of the clustering of the sedentary and activity domains of actigraphy indicate that actigraphy activity and sedentary domains are informative of distinct outcomes, which are only partially and incompletely captured by outcome measures of respiratory questionnaires and in-laboratory lung function and exercise testing. Our findings are consistent with the findings of other studies in which the sedentary domain of actigraphy were predictive of metabolic outcomes and risk of mortality independent of the activity domain.^24,25^

Given that activity monitors can provide objective and reliable data for assessment of health status and disease activity by measuring the real-world physical activity level,^4-7,26,27^ their use has grown significantly in various fields of medicine over the past several years. Actigraphy has become an essential tool in sleep medicine and is increasingly used in the clinical care of patients with sleep and circadian rhythm abnormalities.^28-30^ Through objective measurement of sleep parameters (based on movement) in patient’s home environment, actigraphy provides relevant data with greater external validity compared with self-report and even compared to in-laboratory polysomnography for certain sleep parameters.^31,32^ Actigraphy has also been utilized in telerehabilitation as an alternative to the in-center pulmonary rehabilitation for patients with COPD,^33-35^ or for other rehabilitation programs,^21,22,36,37^ providing measures of physical activity data while the patients are educated and encouraged to participate in daily exercises such as jogging on the treadmill or bicycling. Actigraphy could also allow for high throughput evaluation of large numbers of patients/subjects in both clinical evaluation and clinical research including clinical trials,^38-40^ particularly during the COVID-19 pandemic, which has presented major challenges with in-person and in-laboratory evaluation and testing. Actigraphy is also of particular interest because of the deployment of smartphone applications (App) for tracking daily physical activity, which has been popular due to the common possession of smartphones and, although may exist, they could in principle provide the same measurements as actigraphy does without the need of wearing an exact electronic device.^41,42^ While smartphone App will gradually become more accurate and sustainable from the advancement in software engineering and accessibility of smartphones, actigraphy remains the current method of choice for scaling up standardized physical activity data collection. Altogether, actigraphy could have diverse applications in various medical fields, and thus understanding how it is associated with the current physical activity assessment tools and its limitation are important. Further research with inclusion of larger number of subjects across the entire COPD disease spectrum would be needed to better understand actigraphy application in real-world assessment of physical status of patients.

## Limitations

Our study had several limitations. First, this study had a relatively small sample of 61 participants, which may have diminished the learning power of our machine learning approach for variable selection as well as the statistical power of the final regression modeling. This study was nested in a larger cohort study of the cardiopulmonary health effects of prolonged occupational exposure to SHS, the inclusion and exclusion criteria of which were more stringent and limited our ability to expand our sample size. Nevertheless, our sample size was comparable if not larger relative to other published studies that have characterized physical activity using extensive examination including CPET,^43-45^ and thus provides similar statistical power.

Second, actigraphy data were only collected for five days. In adults, a minimum of 3 to 5 days of accelerometer monitoring is usually considered appropriate to obtain reliable estimates of physical activity.^10^ On the other hand, a longer measurement period of more than 7 days may be desirable to obtain reliable estimates of sedentary behavior since sedentary behavior is more difficult to capture as it might vary more on a day-by-day basis than other activities performed on a higher intensity-level.^46^ However, we aimed to avoid the potential biases that usually occur with variations in level of activity, either increased or decreased, during the weekends by asking the participants to wear the activity monitor for 5 weekdays at the beginning of their working week and the beginning of their day to capture their usual and customary level of activity. In fact, our findings demonstrate a greater number of significant associations for the sedentary domain, suggesting the adequacy of the timeframe within the population studied.

Third, a single activity monitor placed on the waist may not have detected all physical activity since accelerometers have a key limitation in that they are insensitive to certain types of movements, especially non-ambulatory physical activities with arms and/or limbs. However, studies employing multiple accelerometers to increase the accuracy of predicting energy expenditure reported only marginal improvements that would not be justifiable by the increased burden associated with wearing multiple accelerometers.^47,48^

## Conclusion

In conclusion, in this study of a population with early obstructive lung disease or pre-COPD due to prolonged exposure to SHS, we showed that outpatient actigraphy provides “real-world” patient-centered outcomes that objectively inform of patients’ physical activity status. We further showed that the activity and sedentary domains of actigraphy are divergent and provide distinct information that may likely represent the different physiologic processes such as anaerobic and aerobic fitness highlighted by those domains. Furthermore, in this population with pre-COPD, actigraphy measures are not associated with and could not be entirely explained by measures of standardized respiratory questionnaires, lung function, or exercise testing, and thus provide added physical activity measures that could be of value as objective patient-centered outcomes in observational and interventional research studies.

## Data Availability

Data are available upon reasonable request.

## Acknowledgements

We would like to thank Oliver Beech, Patricia Emerson-Healy, Emily Ghio, Liane Tolang, and Charlotte M Hunt for help with performance of cardiopulmonary exercise testing. We also would like to appreciate the contribution of the flight attendants who took time out of their busy schedules to participate as research subjects in this study.

## Contributions

Conceived and designed the experiments: WMG, MA.

Developed study protocols: WCC, WMG, MA.

Collected data: LW, JG, BG, WCC, MN, WMG, MA.

Analyzed and interpreted data: JC, LW, JG, BG, SZ, MA.

Prepared the manuscript: JC, LW, BG, SZ, MA.

Edited the manuscript: JC, LW, JG, BG, SZ, WCC, MN, WMG, MA.

Obtained funding: JG, WMG, MA.

