## Supplemental Appendix for "Actigraphy Informs Distinct Patient-Centered Outcomes in Pre-COPD"

**Running Head:**

Actigraphy in assessment of pre-COPD

**Author List:**

Jianhong Chen, MS ^1,2,3^ *, Lemlem Weldemichael, MD ^1,2,3^ *, Brian Giang, BA ^4^, Jeroen Geerts, MD ^5^, Siyang Zeng, MS ^3,6^, Wendy Czerina Ching, BS ^1,2,3^, Melissa Nishihama, BS ^1,2,3^, Warren M Gold, MD ^1,3^, Mehrdad Arjomandi, MD ^1,2,3^

^1^ Division of Pulmonary, Critical Care, Allergy and Immunology, and Sleep Medicine, Department of Medicine, University of California, San Francisco, California, USA

^2^ Division of Occupational and Environmental Medicine; Department of Medicine, University of California, San Francisco, California, USA

^3^ San Francisco Veterans Affairs Medical Center; San Francisco; California, USA

^4^ George Washington University, School of Medicine, Washington, DC, USA

^5^ Radboud University Medical Center, Nijmegen, the Netherlands

^6^ University of Washington, School of Public Health, Seattle, WA, USA

* These authors contributed equally to this work.

### **SUPPLEMENTAL METHODS**

**DETAILED METHODS**

Study Overview

This was an observational study that was nested in a larger study examining the cardiopulmonary health effects of exposure to SHS in a cohort of nonsmoking subjects with a range of occupational SHS exposure as previously described.^1^ Briefly, between July 2007 and March 2020, we recruited US airline flight crewmembers with a history of occupational exposure to SHS, along with nonsmoker controls without such occupational exposure, who were participating in a larger study of cardiopulmonary health effects of prolonged remote exposure to SHS (ClinicalTrials.gov Identifier: NCT02797275). The participants were characterized by respiratory symptom questionnaires, full pulmonary function testing (PFT), and a maximum effort cardiopulmonary exercise testing (CPET). As part of the actigraphy nested study, beginning February 2014, we invited the participants to wear an activity monitor for eight hours a day for five consecutive days during the week before they come in for in-laboratory evaluation and collected actigraphy data along with a daily diary. The actigraphy data was then analyzed along with the respiratory questionnaires, PFT, and CPET data to examine the association of outpatient actigraphy data with reported physical activity, symptoms, and in-laboratory measures of physical activity.

The University of California San Francisco (UCSF) Institutional Review Board (IRB) and the San Francisco VA Medical Center Committee on Research and Development approved study protocols. Written IRB-approved informed consent and Health Insurance Portability and Accountability Act (HIPAA) were obtained from all study participants. Subjects received monetary compensation for their participation in the study.

Study Population

US airlines crewmembers, including flight attendants and pilots, were eligible to participate if they had worked for ≥5 years in an aircraft. A referent group of ‘sea-level’ subjects who lived in San Francisco Bay area and had never been employed as flight crewmembers were also recruited. Subjects were eligible if they were nonsmokers defined by never-smoking or, in ever smokers, no smoking for ≥20 years and a cumulative history of smoking <20 pack-years. Eligible subjects were excluded if they had a known history of cardiopulmonary disease or recreational drug use including marijuana. All subjects enrolled in the larger cohort were invited to participate in this nested study. For the nested actigraphy study, recruitment began in February 5, 2014 and continued through March 17, 2020.

Physical Activity Monitoring using Actigraphy

Physical activity was monitored using a triaxial accelerometer-based activity monitor (ActiGraph GT3X, Actigraph Corporations, Pensacola, FL). Accelerometer data was recorded in 10-second epochs and in the three “vertical”, “horizontal”, and “perpendicular” axes representing forward, sideways, and upward/downward motion, respectively.

The ActiGraph monitor was initialized to continuously collect data over a period of 5 days. It was then mailed to participants along with a daily diary to keep a log of the time the monitor was worn and the activities the subjects performed during that time. All participants received the ActiGraph monitors at least 7 days prior to the in-laboratory visit, during which data on respiratory symptom questionnaire, PFT and CPET was collected. Participants were instructed to wear the ActiGraph monitor on the waist upon awakening using a waist band provided and to keep it on continuously for at least 8 hours for 5 consecutive days beginning the first day of their work week and beginning the start of their day. The 5-day monitoring was chosen to allow for adequate number of weekday data collection ^2^ while avoiding recording of non-routine rest or activity periods as they may occur during weekends. All participants were carefully instructed on how the device should be positioned.

Actigraphy Data Description

Actigraphy data was processed using the ActiLife software program (Version 6.11.9; ActiGraph LLC) and saved in raw format as GT3X files. The ActiLife software generates a total of 52 variables in the distance, time (activity and sedentary) and energy domains. The list of variables and their definitions are shown in **Table** **S1**. ActiLife outputs use direct measurement of distance, steps, and time for quantifying activity, but quantifying of energy expenditure (EE) requires translation of activity using different prediction models. In this study, we used the Freedson prediction model for translation of activity counts to EE, as it has been commonly used for monitoring of daily non-strenuous physical activity.^3-6^

To further characterize the manner in which sedentary time was accumulated, we created two summary variables in addition to the variables generated by ActiLife: “No. of Sedentary Breaks/Sedentary Time” and “No. of Sedentary Breaks/Avg. Step Count”. (1) A ratio of the number of sedentary breaks over sedentary time (“No. of Sedentary Breaks/Sedentary Time”) was generated to adjust the number of sedentary breaks to the amount of time spent in sedentary activity, which would account for the potential bias of a seemingly increased number of sedentary breaks in subjects who spend more time in sedentary activity; (2) A ratio of the number of sedentary breaks over the average number of steps taken per minute (“No. of Sedentary Breaks/Avg. Step Count”) was generated to adjust the number of sedentary breaks to the same number of steps taken per the total wear time.

Actigraphy Data Processing

Actigraphy data was extracted from the ActiGraph using ActiLife software, which provided a day-by-day summary of collected data. The actigraphy data had a repeated measure design as each subject wore the monitor for 5 consecutive days in the week prior to in-laboratory assessment. Thus, the data was further summarized into 5-day average values (weekly “mean” values) and the highest values of “maximum” values (maximum per epoch) across all 5 days (weekly “highest” values). The actigraphy data was matched against the diary to ascertain the appropriate usage of the monitor, and the data was considered to be acceptable if the subjects wore the monitor for a minimum of 3 days and for greater than 5 hours (300 minutes) per day. The total amount of the time that the monitor was worn was included in the regression models as *total time worn*.

Actigraphy Data Management and Variable Selection

Actigraphy generates a large number of variables, some of which are highly correlated. Given the low number of subjects in our study, we decided to reduce the number of actigraphy variables to increase the robustness of the analysis. To achieve that, we pursued two approaches of variable selection: (1) machine learning approach, and (2) literature-guided approach (**Figure 1**).

In the machine learning approach, we built two models, random forest and lasso regression. From the random forest modeling, we generated a list of top ranked actigraphy variables that were predictive of various questionnaire, PFT, and CPET outcomes by ranking variables based on minimizing prediction error at the splitting of the decision tree nodes. We applied the hyper-parameters of 500 for the number of trees and square root of total dependent variables (13 after rounding in this study) for the number of variables during each split of the decision tree. For lasso regression, we used a similar strategy to generate a list of important variables by utilizing the l_1_-norm penalized terms to force unimportant variables to become zero and setting the hyper-parameter of lambda at 2.009233x10^-2^. To rank the variables in lasso regression, we then compared the magnitude of variables coefficients in various models. Finally, we summarized the actigraphy variables from the two models by counting the number of appearances of the variables in each of the model using the mentioned outcomes and taking the union of the top 10 variables from each list.

In the literature-guided approach, we reviewed the available literature on actigraphy and selected 9 variables (**Table S2**) to represent distance, energy, and activity and sedentary time domains as described below. *Moderate to vigorous physical activity (MVPA)* is a variable representing a category of activity intensity that has been consistently shown to be associated with improved outcomes in many chronic disease states.^7-10^ *Total MVPA* and *Average MVPA/hour* are the two variables selected to represent the total amount of MVPA accumulated over the time worn and the average hourly MVPA, respectively. *Sedentary Time* is a variable representing the amount of time spent in sedentary behavior that has been shown to be associated with increased risk of morbidity and mortality in chronic diseases, often independent of physical activity.^11-15^ *Number of Sedentary breaks*, defined as an interruption in sedentary time where there is change from sedentary to non-sedentary activity, is another time domain that represents the manner in which sedentary time has been accumulated.^16,17^ The occurrence and the pattern by which sedentary time is interrupted has been shown to have important cardiovascular, metabolic, and muscular benefits.^18-20^ *Metabolic equivalents (MET)* and *amount of energy expenditure in kilocalories (Kcals)* are variables representing EE in the energy domain. *Step count* is a distance variable that has been selected as it has been shown to be associated with long-term mortality.^21,22^ Step counts were accumulated on a per epoch basis using accelerometer data collected on axis 1 (“vertical” axis), which best represents energy expenditure during locomotion.^3^ An algorithm present in the device firmware filters out the accelerometer’s baseline noise level to help accurately accumulate the steps-per-epoch.^23^ To evaluate the variability in the number of steps taken throughout the time worn, we selected a maximum and an average value to represent Step Count. *Maximum Step Count* is the maximum number of steps taken per any epoch and *Average Step Count* is the average number of steps taken per minute. Furthermore, Vector Magnitude Counts per Minute (CPM), the vector summation of counts per minute across all three axes (“vertical”, “horizontal” and “perpendicular”), has been included as an additional distance domain variable to represent physical activity in all three dimensions.

The final set of variables was selected based on a combination of the machine learning and literature-guided approaches to provide meaningful variables of highest predictive value for our proposed analysis.

Pulmonary Function Testing

Routine PFT was performed in the seated position using a model Vmax 229 CareFusion (CareFusion Corp., Yorba Linda, CA) and nSpire body plethysmograph (nSpire Health Inc., Longmont, CO). This included measurement of the low-volume curve; spirometry ^24^; lung volume by single breath dilution ^25,26^ and plethysmography ^27^; airway resistance during panting at functional residual capacity (FRC) ^28,29^; and single breath carbon monoxide diffusing capacity. ^30^ Pulmonary function studies were conducted according to the American Thoracic Society (ATS) and European Respiratory Society (ERS) guidelines.^31-36^ Measures of PFT were recorded and percent predicted of normal values were calculated using Crapo predicted formulas.^37-39^

Cardiopulmonary Exercise Testing

Subjects performed physician-supervised, symptom-limited, progressively increasing exercise tests in the supine position on an electromagnetically braked, supine cycle ergometer (Medical Positioning Inc. Kansas City, MO). Beginning December 18, 2015, the exercise tests were done in seated position on an upright cycle ergometer (Corival cpet, 960900, Lode, the Netherlands). Subjects were encouraged to give their best effort, and during testing, were encouraged to continue exercise until a VO_2_ plateau effect on a breath-to-breath analysis of oxygen consumption was visually observed; however, they were advised that they could stop voluntarily at any time they believed they could not continue. We continuously monitored heart rate (HR), systolic and diastolic blood pressure (SBP and DBP), electrocardiogram (ECG), and breath-by-breath gas exchange.

The protocol consisted of 3-min rest, 1-min unloaded (freewheeling) cycling at 60-65 rpm, followed by increasing work rate of 20 to 40 Watts at 2-minute intervals to a maximum tolerated, and 5-min of recovery. Twelve lead ECGs were monitored continuously; ECGs and blood pressure (measured manually by a physician with a cuff) were recorded every 2 min. Oxyhemoglobin saturation (O_2_sat), determined by pulse oximetry, was recorded continuously.

Minute ventilation (V_E_), oxygen uptake (VO_2_) and carbon dioxide output (VCO_2_) were measured breath-by-breath with an open-circuit metabolic cart (model Vmax 229, CareFusion, Yorba Linda, CA). The volumes of the flow meter, mouthpiece, and filter (70 mL x breathing frequency) were subtracted from V_E_ for the V_E_/VCO_2_ calculations. The ratio at which VO_2_ to VCO_2_ was consistently greater or equal to one was used as the Anaerobic Threshold (AT).

Immediately before all tests, the gas analyzers were calibrated using reference gases of known concentrations and the ventilometer was calibrated using a 3-liter syringe (Hans Rudolph, Kansas, MO). The metabolic system was verified using four trained technicians who provided monthly exercise values as biological standards for the laboratory. Percent predicted of normal values were calculated using Wasserman et al. reference formulas.^40^

Respiratory Questionnaires

Patient-reported respiratory symptoms, physical activity, and quality of life assessments were conducted using the COPD Assessment Test (CAT) ^41^, modified Medical Research Council (mMRC) Dyspnea Scale^42^, the Short Form 12-Item Health Survey (SF-12) ^43^, International Physical Activity Questionnaire (IPAQ) ^44^ and Airway Questionnaire 20 (AQ20).^45^

Data Analysis

Distributions of patients actigraphy, respiratory symptoms, lung function and exercise data were visualized and inspected (**Figure S1**). Because the data were not normally distributed, Spearman’s Rank correlation was used for the analysis. We then examined the association of actigraphy distance, energy, and activity and sedentary time domains with respiratory questionnaires, PFT, and CPET outputs after adjustment for age, sex, height, weight, and time worn using hierarchical clustering with Spearman correlation coefficient as the distance metric. We calculated the Spearman correlation coefficient for the final set of actigraphy variable selected and further examined the linear relationships among the variables.

These initial exploratory analyses unveiled two potential statistical challenges of high dimensionality and high collinearity among the actigraphy variables for application of ordinary linear regression modeling. To address these challenges, we employed principal component regression, which combined principal component analysis with adjustment for covariates to reduce the number of dimensions and transformed the original variables into orthogonal principal components (PC) axes. Next, we performed ordinary linear regression using the PC axes as the new predictive actigraphy variables within the model. We later computed the Pearson correlation coefficients between the original actigraphy variables versus the transformed PC axes to help interpret the representation of each PC axis. We used P-values of 0.05 as the usual statistically significant cutoff for all regression models.

### **SUPPLEMENTAL TABLES**

**Table S1- Actigraphy variables and their definition.**

| ACTIGRAPHY VARIABLE | DEFINITION | VARIABLE CODE NAME |
| --- | --- | --- |
| Total Kcals | Measure of EE; total amount of kcals spent during total time worn; a kcal is the amount of energy required to raise the temperature of a liter of water one degree centigrade at sea level |  |
| **Avg. Kcal/hour** | Average kcals spent per hour; total kcal/total time worn (in hours) |  |
| **MET** | Measure of EE; MET is the ratio of a person’s working metabolic rate compared to their resting metabolic rate |  |
| Freedson^3^ Bouts Occurring | The number of Freedson bouts occurring during total time worn |  |
| Freedson^3^ Bouts Starting | The number of Freedson bouts starting during the total time worn |  |
| Freedson^3^ Bouts Ending | The number of Freedson bouts ending during the total time worn |  |
| Total Time of Freedson^3^ Bouts | Total time (in min) of Freedson bouts occurring during total time worn |  |
| Total Counts of Freedson^3^ Bouts | Total activity counts of Freedson bouts occurring during total time worn |  |
| Sedentary Bouts Occurring | The number of sedentary bouts occurring during total time worn |  |
| Sedentary Bouts Starting | The number of sedentary bouts starting during total time worn |  |
| Sedentary Bouts Ending | The number of Freedson bouts ending during the total time worn |  |
| Total Time of Sedentary Bouts | Total time (in min) of sedentary bouts occurring during total time worn |  |
| **No. of Sedentary Breaks** | The number of sedentary breaks occurring during total time worn; sedentary break is an interruption from sedentary behavior |  |
| No. of Sedentary Breaks Starting | The number of sedentary breaks starting during total time worn |  |
| No. of Sedentary Breaks Ending | The number of sedentary breaks ending during total time worn |  |
| Total Time of Sedentary Breaks | Total time (in min) of sedentary breaks occurring during total time worn |  |
| **Sedentary Time** | Length of time (in min) in sedentary behavior |  |
| Light Time | Length of time (in min) in light PA |  |
| Moderate Time | Length of time (in min) in moderate PA |  |
| Vigorous Time | Length of time (in min) in vigorous PA |  |
| Very Vigorous Time | Length of time (in min) in very vigorous PA |  |
| % in Sedentary Time | Percent of time in sedentary behavior |  |
| % in Light Time | Percent of time in light PA |  |
| % in Moderate Time | Percent of time in moderate PA |  |
| % in Vigorous Time | Percent of time in vigorous PA |  |
| % in Very Vigorous Time | Percent of time in very vigorous PA |  |
| **Total MVPA** | Total time (in min) in MVPA |  |
| % in MVPA | Percent of time in MVPA |  |
| **Average MVPA/hour** | Average amount of MVPA (in min) per hour |  |
| Axis 1^#^ Counts | Sum of counts for axis 1 (Y-axis) |  |
| Axis 2^#^ Counts | Sum of counts for axis 2 (X-axis) |  |
| Axis 3^#^ Counts | Sum of counts for axis 3 (Z-axis) |  |
| Axis 1^#^ Average Counts | Average of counts for axis 1 (Y-axis) |  |
| Axis 2^#^ Average Counts | Average of counts for axis 2 (X-axis) |  |
| Axis 3^#^ Average Counts | Average of counts for axis 3 (Z-axis) |  |
| Axis 1^#^ Max Counts | Maximum count value for axis 1 (Y-axis); max = max count / epoch |  |
| Axis 2^#^ Max Counts | Maximum count value for aAxis 2; max = max count /epoch (X-axis) |  |
| Axis 3^#^ Max Counts | Maximum count value for aAxis 3; max = max count /epoch (Z-axis) |  |
| Axis 1^#^ CPM | Counts per minute for axis 1 (Y-axis) |  |
| Axis 2^#^ CPM | Counts per minute for axis 2 (X-axis) |  |
| Axis 3^#^ CPM | Counts per minute for axis 3 (Z-axis) |  |
| Vector Magnitude Counts | Vector magnitude of all 3 axes |  |
| Vector Magnitude Avg. Counts | Average vector magnitude of all 3 axes |  |
| Vector Magnitude Max. Counts | Maximum vector magnitude of all 3 axes; max = max count /epoch |  |
| **Vector Magnitude CPM** | Vector magnitude counts per minute |  |
| Total Steps Counts^+^ | Sum of step counts during total time worn |  |
| Avg. Step Count/Epoch | Average number of steps per every epoch |  |
| **Max. Step Count** | Maximum step counts; maximum of daily maximum step count per/epoch |  |
| **Avg. Step Count** | Average number of steps taken per min |  |
| Number of Epochs | Number of epochs, epoch = 10s period of time |  |
| Total Time Worn | Length of scored time (in min) |  |
| Date | Date ActiGraph monitor worn |  |

Footnote: Names and definition of 52 actigraphy variable outputs by the ActiLife software.

Bolded variables have been selected as the most informative actigraphy variables using the literature-guided approach. Refer Table S2 below for more detailed definition and references. Data has been collected in 10sec epochs.

**^#^**Axis 1 (Y-Axis) – “vertical” (forward) axis activity acceleration data

**^#^**Axis 2 (X-Axis) – “horizontal” (sideways) axis activity acceleration data

**^#^**Axis 3 (Z-Axis) – “perpendicular” (upward/downward) axis activity acceleration data

**^+^**Step Counts are accumulated on a per epoch basis and are based on accelometer data collected in the **vertical** axis

Abbreviations- EE: energy expenditure; MET: metabolic equivalent; CPM: counts per minute; MVPA: moderate to vigorous physical activity

**Table S2- Actigraphy variable selection using literature-guided approach.**

| **Variable name** | **Variable definition** | **Literature citation** | **Reference** |
| --- | --- | --- | --- |
| Total MVPA | Physical activity that involves energy expenditure above 3.0 METs; time (in min) when CPM ≥1952 | Saint-Maurice, Troiano et al. 2018, Kraus, Powell et al. 2019, Fini, Bernhardt et al. 2020, Lee, Walker et al. 2020 | ^7-10^ |
| Avg. MVPA/hour | Average time spent in MVPA; total MVPA (in min)/total time worn (in hours) | Saint-Maurice, Troiano et al. 2018, Kraus, Powell et al. 2019, Fini, Bernhardt et al. 2020, Lee, Walker et al. 2020 | ^7-10^ |
| Sedentary Time | Time spent in PA that does not increase EE substantially above the resting level (1.0 - 1.5 METs); time (in min) when CPM < 100 | Healy, Dunstan et al. 2008, Pate, O'Neill et al. 2008, Proper, Singh et al. 2011, Thorp, Owen et al. 2011, Koster, Caserotti et al. 2012, Patterson, McNamara et al. 2018 | ^11-15,46^ |
| No. of Sedentary Breaks | Activity count change from <100 to ≥ 100 counts. A break only occurs if the change in count is sustained for more than 2 min | Bey and Hamilton 2003, Hamilton, Hamilton et al. 2007, Healy, Dunstan et al. 2008, Altenburg and Chinapaw 2015, Tremblay, Aubert et al. 2017 | ^16-20^ |
| Max. Step Count | Maximum of the daily maximum number of steps taken per epoch (10 sec) | Dwyer, Pezic et al. 2015, Kraus, Yates et al. 2018 | ^21,22^ |
| Avg. Step Count | Average number of steps taken per min; total step count/total time worn (in min) | Dwyer, Pezic et al. 2015, Kraus, Yates et al. 2018 | ^21,22^ |
| Vector Magnitude CPM | Vector summation of CPM across all 3 axes (vertical, horizontal and perpendicular) | Butte, Wong et al. 2014, Kim, Lee et al. 2014, Aadland and Ylvisåker 2015 | ^47-49^ |
| Avg. Kcal/hour | Measure of EE; a kcal is the amount of energy required to raise the temperature of a liter of water one degree centigrade at sea level; total kcal/total time worn (in hours) | Midorikawa, Tanaka et al. 2007, Plasqui and Westerterp 2007, Ainsworth, Haskell et al. 2011 | ^50-52^ |
| MET | Measure of EE; MET is the ratio of a person’s working metabolic rate compared to their resting metabolic rate; sedentary behavior 1.0-1.5 METs, light- intensity PA 1.6-2.9 METs, moderate-intensity PA ≥3.0 to <6.0 METs, vigorous-intensity ≥6.0 to <9.0 METs and very vigorous PA is ≥9.0 METs | Plasqui and Westerterp 2007, Lee, Djoussé et al. 2010, Ainsworth, Haskell et al. 2011 | ^50,51,53^ |

Footnote: Definitions and literature citations for actigraphy variables selected using the literature-guided approach.

Abbreviations- MVPA: moderate to vigorous physical activity; MET: metabolic equivalent; CPM: counts per minute; PA: physical activity; EE: energy expenditure.

### **SUPLLEMENTAL FIGURES**

**Figure S1- Spearman’s Rank correlation among selected actigraphy variables.** The matrix plot shows the interactions among actigraphy variables via scatter plot (bottom left), distribution of each individual variables (diagonal), and the Spearman correlation Coefficients (top right).

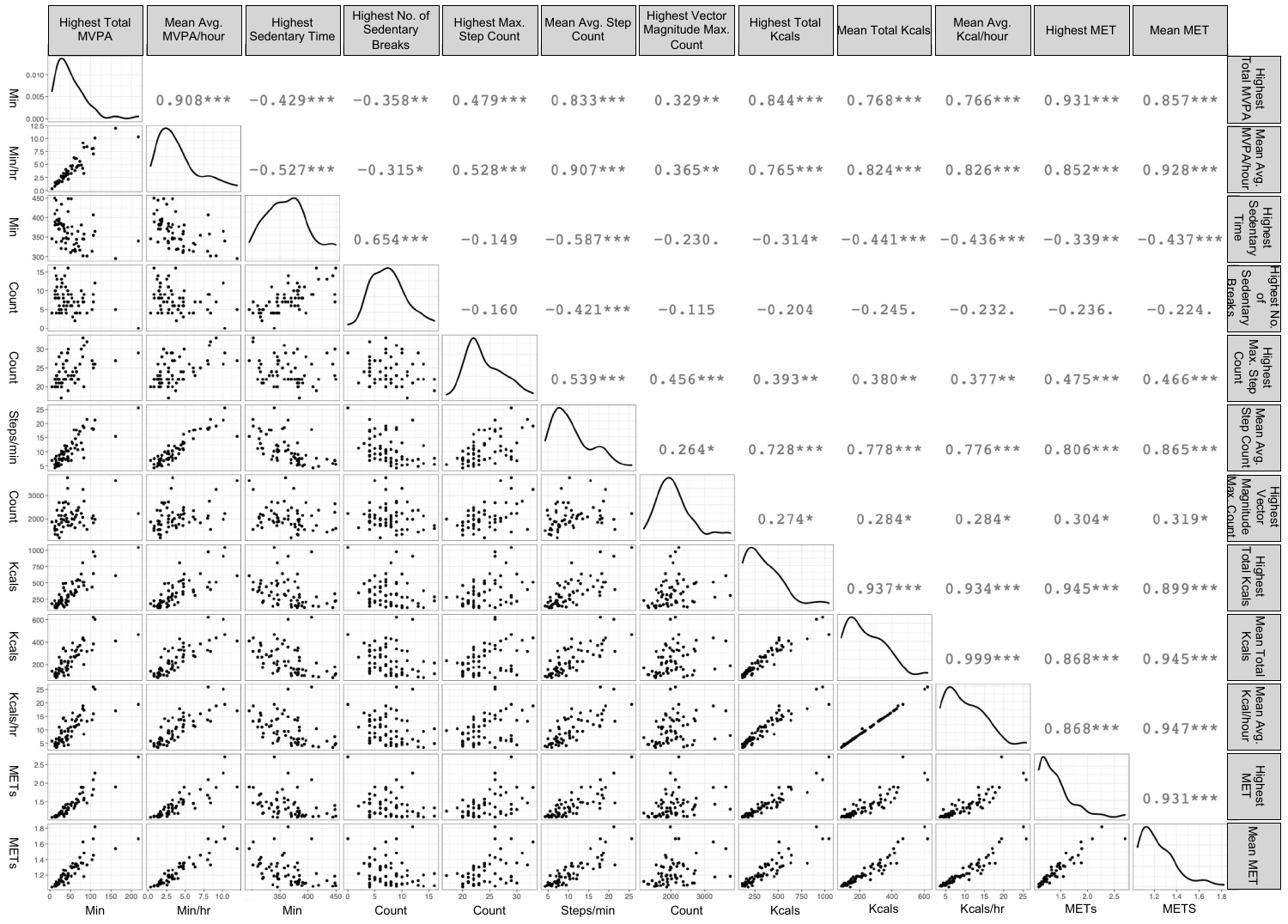

**Figure S2- Random Forest and Lasso regression models for actigraphy variable selection**. (A) Random forest modeling was for each of the respiratory questionnaire, PFT, and CPET outcomes over actigraphy variables, and the variables were ranked based on the number of appearances from each model. Random Forest hyper parameter setting: No. Tree = 500, No. features = 15, minimum node size = 5. (B) Similar to random forest modeling, lasso regression was applied and the same ranking by number of appearances from each model was performed. Lasso Regression hyper parameter setting: Lambda = 2.009233x10^-2^. Finally, the final machine learning selected variables are derived by taking the union of the top 10 variables from Fig. S2A and Fig. S2B.

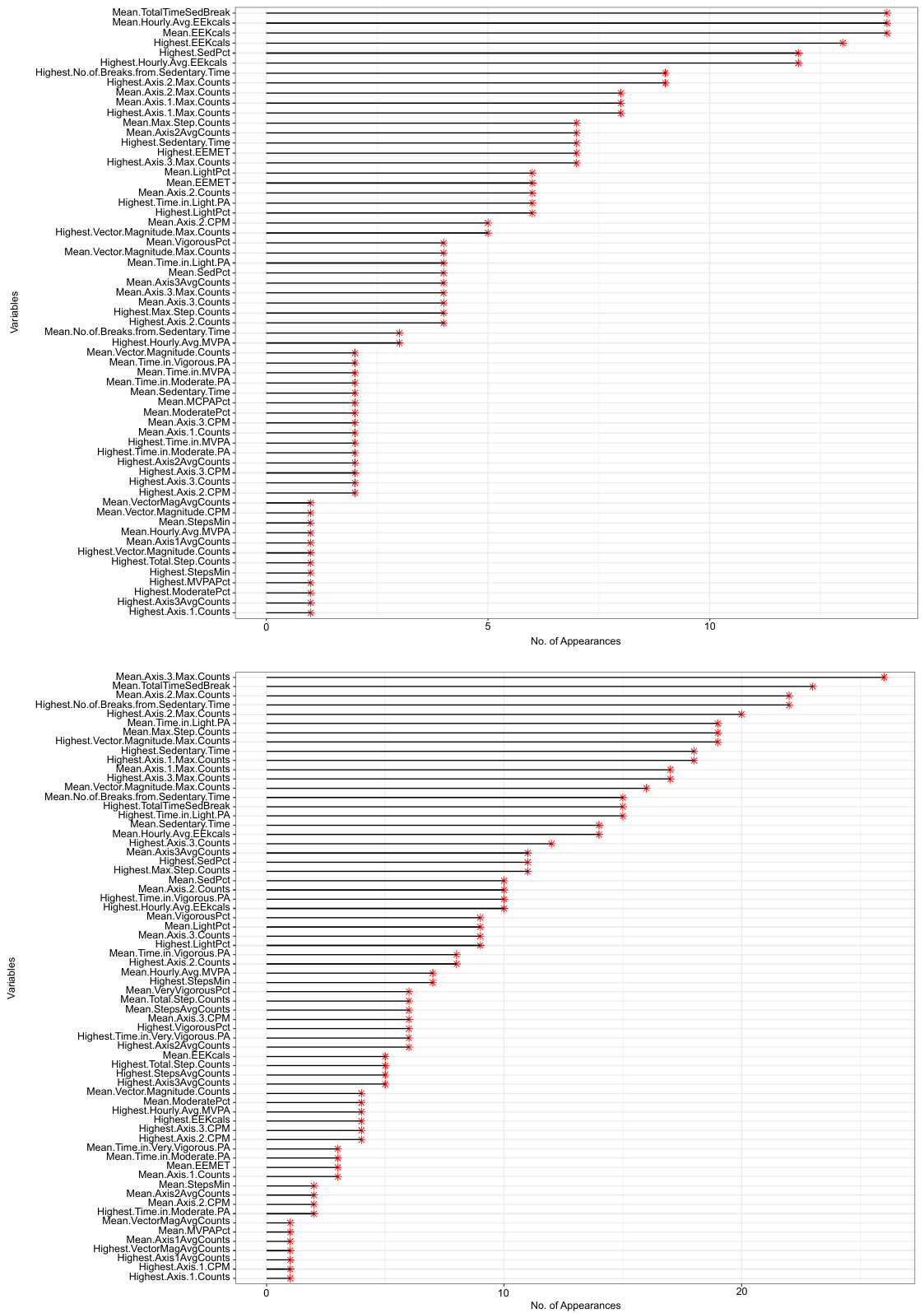

**Figure S3 – Hierarchical clustering analysis and heatmap representation of all actigraphy variables with questionnaire, PFT and CPET measures.** We performed hierarchical clustering using Spearman correlation coefficients as the distance metric. The clustered relationships for the variables were present as the dendrograms on the top of the heat map. Two distinct clusters of actigraphy variables were generated from the three domains (time, distance, energy). The two clusters were based on activity-related (such as distance walked, amount of energy spent, etc.) and sedentary-related (such as sedentary time, break, etc.) respectively. Each row represents one subjects. Grey boxes mean the values were missing.

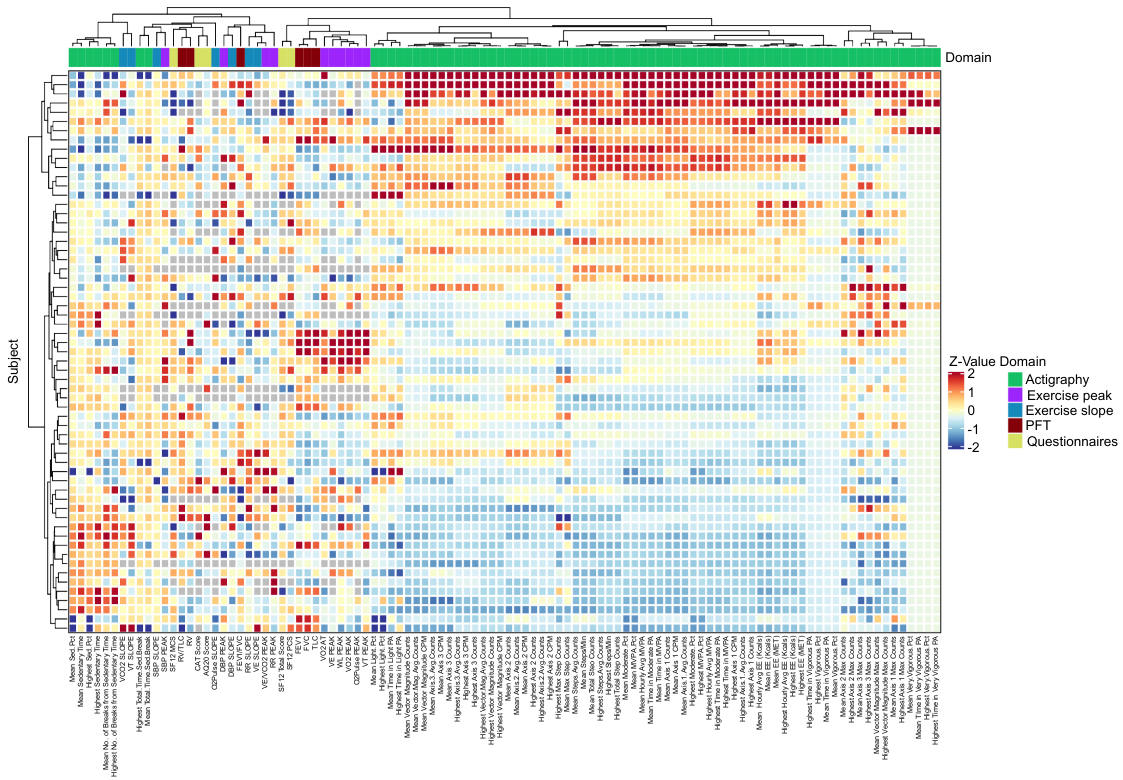
